# Pervasive correlations between causal disease effects of proximal SNPs vary with functional annotations and implicate stabilizing selection

**DOI:** 10.1101/2023.12.04.23299391

**Authors:** Martin Jinye Zhang, Arun Durvasula, Colby Chiang, Evan M. Koch, Benjamin J. Strober, Huwenbo Shi, Alison R. Barton, Samuel S. Kim, Omer Weissbrod, Po-Ru Loh, Steven Gazal, Shamil Sunyaev, Alkes L. Price

**Author notes:** These authors contributed equally (Arun Durvasula, Colby Chiang).

## Abstract

The genetic architecture of human diseases and complex traits has been extensively studied, but little is known about the relationship of causal disease effect sizes between proximal SNPs, which have largely been assumed to be independent. We introduce a new method, LD SNP-pair effect correlation regression (LDSPEC), to estimate the correlation of causal disease effect sizes of derived alleles between proximal SNPs, depending on their allele frequencies, LD, and functional annotations; LDSPEC produced robust estimates in simulations across various genetic architectures. We applied LDSPEC to 70 diseases and complex traits from the UK Biobank (average *N*=306K), meta-analyzing results across diseases/traits. We detected significantly nonzero effect correlations for proximal SNP pairs (e.g., −0.37 *±*0.09 for low-frequency positive-LD 0-100bp SNP pairs) that decayed with distance (e.g., −0.07 *±*0.01 for low-frequency positive-LD 1-10kb), varied with allele frequency (e.g., −0.15 *±*0.04 for common positive-LD 0-100bp), and varied with LD between SNPs (e.g., +0.12 *±*0.05 for common negative-LD 0-100bp) (because we consider derived alleles, positive-LD and negative-LD SNP pairs may yield very different results). We further determined that SNP pairs with shared functions had stronger effect correlations that spanned longer genomic distances, e.g., −0.37 *±*0.08 for low-frequency positive-LD same-gene promoter SNP pairs (average genomic distance of 47kb (due to alternative splicing)) and −0.32 *±*0.04 for low-frequency positive-LD H3K27ac 0-1kb SNP pairs. Consequently, SNP-heritability estimates were substantially smaller than estimates of the sum of causal effect size variances across all SNPs (ratio of 0.87 *±*0.02 across diseases/traits), particularly for certain functional annotations (e.g., 0.78 *±*0.01 for common Super enhancer SNPs)—even though these quantities are widely assumed to be equal. We recapitulated our findings via forward simulations with an evolutionary model involving stabilizing selection, implicating the action of linkage masking, whereby haplotypes containing linked SNPs with opposite effects on disease have reduced effects on fitness and escape negative selection.

## Introduction

Inferring the genome-wide distribution of causal genetic effects has yielded rich insights into the polygenic architecture of human diseases and complex traits^1–21^. However, virtually all published studies of disease and complex trait architectures assume that nearby SNPs have independent causal effects on disease^1–21^—an assumption that warrants careful scrutiny. Correlated effects may arise due to natural selection^22–28^, e.g., due to linkage masking, whereby haplotypes containing linked SNPs with opposite effects on disease have a reduced aggregate effect on fitness and escape negative selection^23,28^; correlated effects have also been reported in studies of rare coding variants (concordant effects^29–34^) and model organisms (concordant^35^ or opposite^35,36^ effects). Despite these findings, SNP-pair effect correlations have yet to be systematically investigated in genome-wide data.

Here, we propose a method, linkage disequilibrium SNP-pair effect correlation regression (LDSPEC), to estimate correlations of standardized derived allele causal disease effect sizes for pairs of proximal SNPs, depending on their minor allele frequency (MAF), LD, and functional annotations. Roughly, LDSPEC determines that a SNP-pair annotation has positive (resp. negative) correlation of causal effect sizes (of derived alleles) if SNPs with concordant signed LD to pairs of SNPs in the SNP-pair annotation have higher (resp. lower) *χ*^2^ statistics than SNPs with discordant signed LD. We performed extensive simulations with real genotypes to show that LDSPEC is well-calibrated in null simulations and produces attenuated estimates of nonzero SNP-pair effect correlations in causal simulations. We applied LDSPEC to 70 UK Biobank diseases and complex traits^37^ (*N*=306K), estimating effect correlations for common (MAF ≥ 5%) positive-LD, common negative-LD, low-frequency (0.5% ≤ MAF*<*5%) positive-LD, and low-frequency negative-LD SNP pairs depending on their functional annotations. We note that because we consider derived alleles, positive-LD and negative-LD SNP pairs differ in a way that is not arbitrary and may yield very different results. We recapitulated our findings via forward simulations with an evolutionary model involving stabilizing selection^38,39^.

We note that this study expands upon an unpublished preprint^40^, which contained key ideas and derivations and detected SNP-pair effect correlations for extremely-short-range SNP pairs (0-100bp) that varied with LD; here, we introduce improved methodology, analyze functional SNP-pair annotations, identify SNP-pair effect correlations at longer distances, and perform evolutionary forward simulations to interpret our findings.

## Results

### Overview of methods

LDSPEC estimates the signed correlation of standardized derived allele causal disease effect sizes across SNP pairs in a given SNP-pair annotation, e.g., set of 0-100bp SNP pairs. The method adopts and improves upon key ideas and derivations from a recent preprint^40^ (see Discussion). In detail, for a SNP-pair annotation defined by a set of SNP pairs *G*, LDSPEC estimates the SNP-pair effect correlation

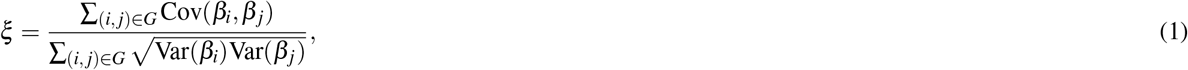

where *β*_*i*_,*β* _*j*_ denote standardized derived allele causal disease effect sizes of SNPs *i, j* (i.e., number of standard deviations increase in phenotype per 1 standard deviation increase in genotype) under a random-effects model, Var(*β*_*i*_),Var(*β* _*j*_) denote expected per-SNP heritabilities, and Cov(*β*_*i*_, *β*_*j*_) denotes expected per-SNP-pair effect covariance. We note that previous work has broadly assumed that causal effects are independent^1–21^ (implying *ξ* = 0), but LDSPEC challenges this assumption. To assess correlations specific to the SNP-pair annotation, LDSPEC also estimates the excess SNP-pair effect correlation *ξ*^∗^, defined as the difference between *ξ* and its expected value across distance-matched SNP pairs. To assess the impact of SNP-pair effect correlations on SNP-heritability, LDSPEC separately estimates genome-wide SNP-heritability and the sum of causal effect size variances across SNPs (SCV = ∑_*i*_ Var(*β*_*i*_)); the two quantities may be different when causal effects are not independent (as assumed in previous work^1–21^).

LDSPEC relies on the fact that the *χ*^2^ association statistic for a given SNP includes the effects of all SNPs tagged by that SNP^4,41^. Methods for analyzing single-SNP annotations^5^ determine that a single-SNP annotation is enriched for heritability if SNPs with higher LD to SNPs in the single-SNP annotation have higher *χ*^2^ statistics than SNPs with low LD to SNPs in the single-SNP annotation. LDSPEC further determines that a SNP-pair annotation has a positive (resp. negative) correlation of causal effect sizes (of derived alleles) if SNPs with concordant signed LD to SNP pairs in the SNP-pair annotation have higher (resp. lower) *χ*^2^ statistics than SNPs with discordant signed LD to SNP pairs in the SNP-pair annotation.

In detail, under a polygenic model^1^, the expected *χ*^2^ of SNP *i* can be written as

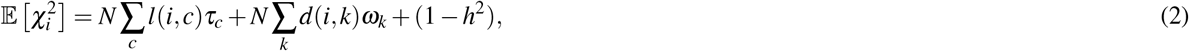

where *N* is the GWAS sample size, *l*(*i, c*) is the LD score of SNP *i* and single-SNP annotation *c* (ref.^5,8^) (defined as 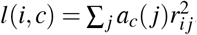, where *a*_*c*_(*j*) is the value of single-SNP annotation *c* for SNP *j* and *r*_*i j*_ is the signed LD between SNPs *i, j*), *τ*_*c*_ denotes the contribution of single-SNP annotation *c* to per-SNP heritability (ref.^5,8^), *d*(*i, k*) is the directional LD score of SNP *i* and SNP-pair annotation *k* (defined as 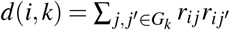 where *G*_*k*_ is the set of SNP pairs in SNP-pair annotation *k*), *ω*_*k*_ denotes the contribution of SNP-pair annotation *k* to per-SNP-pair effect covariance, and *h*^2^ denotes disease/trait SNP-heritability. The last term (1− *h*^2^) is different from 1 in the analogous LDSC equation^4,5^ because LDSC uses an external LD reference panel while our method uses in-sample LD to avoid challenges that arise from the use of inaccurate LD reference panels^11,42,43^ (Methods); we also use a larger 10Mb LD window compared to the 1Mb window commonly used in LDSC^4,5^. Equation (2) allows us to estimate *τ*_*c*_ and *ω*_*k*_ via multivariate linear regression of 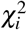 on *l*(*i, c*) and *d*(*i, k*), and we can further estimate quantities such as *ξ* and *ξ*^∗^ based on estimates of *τ*_*c*_ and *ω*_*k*_. We employ regression weights to account for dependency between regression SNPs and heteroskedasticity, and estimate standard errors via genomic block-jackknife, analogous to previous work^4,5^. Further details are provided in the Methods section and Supplementary Note; we have publicly released open-source software implementing LDSPEC (see Code availability).

We applied LDSPEC to 70 well-powered diseases and complex traits from the UK Biobank^37^ (z-score >5 for nonzero SNP-heritability; average *N*=305,646 unrelated “in.white.British.ancestry.subset” individuals, a previously-defined subset of UK Biobank participants who self-reported White British ethnicity and had very similar genetic ancestry based on principal component analysis), including 29 independent diseases/traits (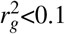, average *N*=298,430) (Supplementary Table 1; see Data availability). We considered 14,820,648 imputed SNPs (version “imp_v3” from ref.^37^, MAF ≥ 0.1%, INFO score^44^ ≥ 0.6, ref.^11,43^). We analyzed 165 single-SNP annotations, including 163 baseline-LF annotations^11^ and 2 annotations for deleterious coding SNPs (common and low-frequency SNPs with CADD pathogenicity score^45^ >20, resp.) (Supplementary Tables 2,3). We refer to the heritability model defined by the 165 single-SNP annotations as the “baseline” model. We further constructed a “baseline-SP” model including, in addition to the 165 single-SNP annotations, 136 SNP-pair annotations obtained by stratifying 34 main SNP-pair annotations by MAF (common or low-frequency) and LD (positive or negative): 3 proximity-based annotations (0-100bp, 100bp-1kb, 1-10kb), 5 gene-based annotations (e.g., same-gene promoter SNP pairs), 7 functional 0-100bp annotations, and 19 functional 0-1kb annotations (e.g., pairs of H3K27ac SNPs with distances 0-100bp) (Table 1, Supplementary Tables 4,5). The functional SNP-pair annotations were constructed from 38 binary baseline single-SNP functional annotations, subject to a requirement to yield at least 1 million SNP pairs (this requirement is more difficult to satisfy for 0-100bp annotations, implying a smaller number of functional 0-100bp annotations retained). We excluded SNP-pair annotations involving one common SNP and one low-frequency SNP, because these SNP pairs had low levels of LD, limiting the informativeness of directional LD scores (Methods). We have publicly released all SNP annotations and LDSPEC output from this study (see Data availability).

**Table 1.** Main SNP-pair annotations. We report the name, number of SNP pairs, and average distance, for each of 34 SNP-pair annotations in the baseline-SP model (136 SNP-pair annotations when counting common positive-LD, low-frequency positive-LD, common negative-LD, and low-frequency negative-LD SNP-pair annotations separately): 3 proximity-based annotations, 5 gene-based annotations, 7 functional 0-100bp annotations, and 19 functional 0-1kb annotations. Further details are provided in Supplementary Table 4.

|  | Number of SNP pairs | Average distance |
| --- | --- | --- |
| Proximal 0-100bp | 3.5M | 47bp |
| Proximal 100bp-1kb | 27M | 546bp |
| Proximal 1-10kb | 253M | 5.4kb |
| Same-exon | 0.81M | 3.6kb |
| Same-gene exonic | 1.8M | 53kb |
| Same-gene promoter | 1.2M | 46kb |
| Same-protein-domain | 0.19M | 47kb |
| Same-gene | 1889M | 390kb |
| H3K27ac-100 | 1.4M | 46bp |
| H3K27ac (PGC2)-100 | 0.92M | 46bp |
| H3K4me1-100 | 1.4M | 46bp |
| Intron-100 | 1.3M | 46bp |
| Repressed-100 | 1.6M | 45bp |
| Super enhancer-100 | 0.61M | 46bp |
| Transcribed-100 | 1.1M | 44bp |
| DGF-1k | 1.2M | 387bp |
| DHS-1k | 1.9M | 383bp |
| DHS peaks-1k | 0.91M | 366bp |
| Enhancer-1k | 0.73M | 418bp |
| Fetal DHS-1k | 0.84M | 349bp |
| H3K27ac-1k | 11M | 483bp |
| H3K27ac (PGC2)-1k | 7.2M | 469bp |
| H3K4me1-1k | 10M | 466bp |
| H3K4me1 peaks-1k | 2.1M | 432bp |
| H3K4me3-1k | 2.7M | 436bp |
| H3K9ac-1k | 2.5M | 441bp |
| Intron-1k | 11M | 487bp |
| Promoter-1k | 1.3M | 469bp |
| Repressed-1k | 10M | 466bp |
| Super enhancer-1k | 5.3M | 487bp |
| TFBS-1k | 1.9M | 388bp |
| Transcribed-1k | 6.8M | 458bp |
| Super enhancer (Vahedi)-1k | 0.59M | 485bp |
| Typical enhancer-1k | 0.65M | 475bp |

### Simulations assessing calibration and power

We performed null simulations (heritable traits with zero SNP-pair effect correlations) and causal simulations (heritable traits with nonzero SNP-pair effect correlations). We used the same UK Biobank genotype data (*N*=337,426) and restricted to chromosome 1 SNPs (*M*=1,161,341) for computational tractability (analogous to ref.^8,11^). In our primary simulations, SCV was set to 0.5 (similar to previous work^11^; SNP-heritability was slightly different from SCV when SNP-pair effect correlations were nonzero), causal SNP proportion was set to 0.2 (similar to previous work^11^), LD-dependent and MAF-dependent genetic architectures were specified based on previous work^8,11^, and functional enrichment was simulated by assigning a positive *τ* to the common Super enhancer (Hnisz) single-SNP annotation; other settings were also evaluated. True simulated values of nonzero SNP-pair effect correlations for SNP-pair annotations in causal simulations are described below, and generative model parameters for all simulations are provided in Supplementary Table 7. Results were obtained by running LDSPEC with the baseline-SP model. We assessed bias (in null and causal simulations) and power (in causal simulations) using mean estimates and empirical SEs across 50 simulation replicates (empirical SE = empirical 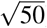), and assessed calibration (in null and causal simulations) by comparing average jackknife SE (across 50 simulation replicates) to empirical SD; we note that aggregating 50 simulation replicates reduces the empirical SE, analogous to meta-analyzing 29 independent diseases/traits in real data. Further details are provided in the Methods section.

We first performed null simulations, simulating heritable traits with functional enrichment but zero SNP-pair effect correlations for all SNP-pair annotations. We reached 6 main conclusions. First, estimates of SNP-pair effect correlation (*ξ*) were approximately unbiased, with no significant bias for all 136 SNP-pair annotations (*P*>0.05/136) (Figure 1a and Supplementary Table 8); furthermore, we did not observe a trend towards negative *ξ* for positive-LD SNP pairs or positive *ξ* for negative-LD SNP pairs. Second, estimates of excess SNP-pair effect correlation (*ξ*^∗^) were approximately unbiased, with no significant bias for all 136 SNP-pair annotations (*P*>0.05/136) (Supplementary Figure 1a). Third, estimates of the contribution of a SNP-pair annotation to per-SNP-pair effect covariance (*ω*) were approximately unbiased, with no significant bias for all 136 SNP-pair annotations (*P*>0.05/136) (Supplementary Figure 1a). Fourth, estimates of the contribution of a single-SNP annotation to per-SNP heritability (*τ*), total SNP-heritability, and heritability enrichment were approximately unbiased, analogous to previous work^5,8,11^ (Supplementary Figure 2a). Fifth, distinct from estimates of total SNP-heritability, estimates of the sum of causal effect size variances across all SNPs (total SCV), as well as total heritability shrinkage (total SCV divided by total SNP-heritability) were approximately unbiased (Supplementary Figure 2a). Sixth, jackknife standard errors for all quantities were well-calibrated (Supplementary Figure 3a).

**Figure 1.**
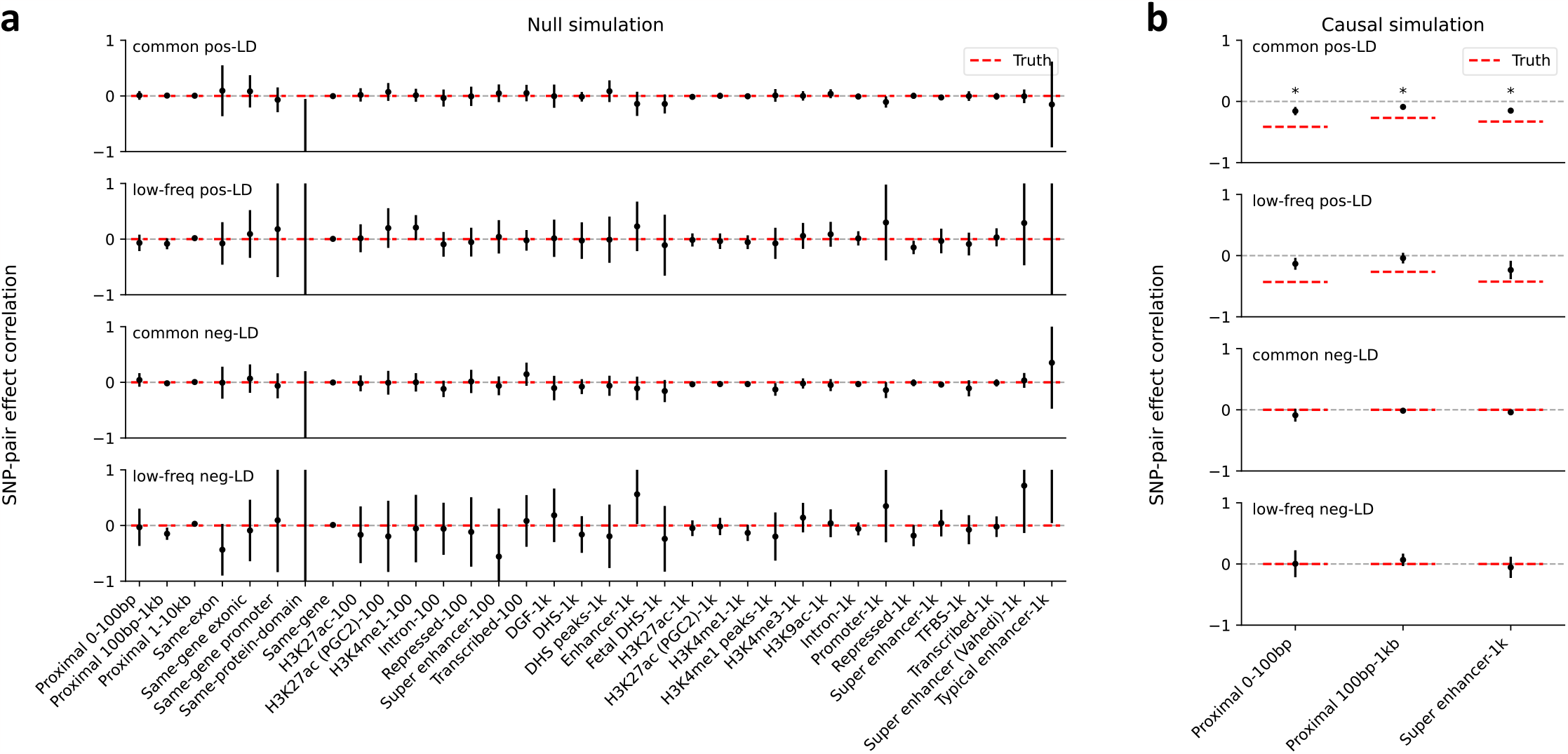
Estimates of SNP-pair effect correlations in null and causal simulations. **(a)** Null simulations with zero SNP-pair effect correlation. We report estimates of SNP-pair effect correlation (*ξ*) for the 136 SNP-pair annotations in the baseline-SP model. Error bars denote 95% confidence intervals around the mean of 50 simulation replicates; “*” denotes statistical significance after multiple testing correction (*P*<0.05/136). Numerical results are reported in Supplementary Table 8. **(b)**Causal simulations with negative SNP-pair effect correlations for a subset of positive-LD SNP-pair annotations. We report estimates of SNP-pair effect correlation (*ξ*) for the 6 causal positive-LD SNP-pair annotations simulated to have negative contribution to per-SNP-pair effect covariance (*ω*) and the corresponding 6 non-causal negative-LD SNP-pair annotations. Error bars denote 95% confidence intervals around the mean of 50 simulation replicates; “*” denotes statistical significance after multiple testing correction (*P*<0.05/136). Red dashed lines denote true simulated values. Numerical results are reported in Supplementary Table 9.

We next performed causal simulations, simulating heritable traits with functional enrichment and nonzero SNP-pair effect correlation for a subset of SNP-pair annotations. To mimic results in real data (see below), we specified negative contributions to per-SNP-pair effect covariance (*ω*) for 6 positive-LD SNP-pair annotations (common and low-frequency 0-100bp, 100bp-1kb, and super-enhancer 0-1kb; zero *ω* for the 6 corresponding negative-LD SNP-pair annotations; Supplementary Table 7); other SNP-pair annotations that overlap the 6 causal SNP-pair annotations are expected to have nonzero SNP-pair effect correlation (*ξ*). We reached 6 main conclusions. First, estimates of SNP-pair effect correlation (*ξ*) were significantly negative for all 3 causal common positive-LD SNP-pair annotations (*P*<0.05/136), non-significantly negative for all 3 causal low-frequency positive-LD SNP-pair annotations (*P*>0.05/136), and attenuated towards 0 for all 6 causal SNP-pair annotations (Figure 1b and Supplementary Table 9); estimates were non-significant for the 6 corresponding negative-LD SNP-pair annotations (*P*>0.05/136), consistent with their zero simulated *ξ*. 10 of the remaining 62 non-causal positive-LD SNP-pair annotations had significantly negative estimates (*P*<0.05/136), as expected due to overlap with the 6 causal positive-LD SNP-pair annotations (Supplementary Figure 1b). 1 negative-LD SNP-pair annotations had a slightly but significantly positive estimate (common negative-LD 1-10kb, 0.016 *±* 0.004) (*P*<0.05/136) (Supplementary Figure 1b), suggesting a slight bias (perhaps due to collinearity of directional LD scores between SNP-pair annotations (Supplementary Table 6)); we believe that this should not impact our interpretation of results in real data, as the magnitude of *ξ* estimates was much larger in real data (see below) and LDSPEC produced unbiased estimates in null simulations. Second, estimates of excess SNP-pair effect correlation (*ξ*^∗^) were significantly negative for the two SNP-pair annotations that were simulated to have negative *ξ*^∗^ (common and low-frequency positive-LD super-enhancer 0-1kb) (*P*<0.05/136) (Supplementary Figure 1b). 4 other positive-LD functional SNP-pair annotations also had significantly negative *ξ*^∗^ estimates (*P*<0.05/136), as expected due to overlap with the causal SNP-pair annotations (Supplementary Figure 1b). 1 negative-LD functional SNP-pair annotation had a slightly but significantly positive estimate (common negative-LD intron 0-1kb, 0.061 *±* 0.014) (*P*<0.05/136) (Supplementary Figure 1b), suggesting a slight bias (perhaps due to collinearity of directional LD scores between SNP-pair annotations (Supplementary Table 6), analogous to the *ξ* estimates above); we believe that this should not impact our interpretation of results in real data, as we detected substantially more significantly positive *ξ*^∗^ estimates for negative-LD functional SNP-pair annotations with larger magnitudes in real data (see below) and LDSPEC produced unbiased estimates in null simulations. Third, estimates of the contribution of a SNP-pair annotation to per-SNP-pair effect covariance (*ω*) were significantly negative for 1 of 6 causal SNP-pair annotations (common Super enhancer 0-1kb) (*P*<0.05/136) but non-significant and attenuated towards 0 for the other 5 (*P*>0.05/136) (Supplementary Figure 1b). 4 of the 130 non-causal SNP-pair annotations also had significantly nonzero estimates (low-frequency positive-LD 1-10kb, common negative-LD 1-10kb, common positive-LD intron 0-1kb, common negative-LD intron 0-1kb) (*P*<0.05/136) (Supplementary Figure 1b) (perhaps due to the collinearity of directional LD scores between SNP-pair annotations (Supplementary Table 6), analogous to the *ξ* estimates above); we believe that this should not impact our interpretation of results in real data, as analyses of real data primarily focused on *ξ* estimates (see below) and LDSPEC produced unbiased *ω* estimates in null simulations. Fourth, estimates of the contribution of a single-SNP annotation to per-SNP heritability (*τ*) were attenuated towards 0 (7.4 *×*10^−7^ *±* 4.3 *×*10^−8^, true value 1.9 *×*10^−6^ for the common Super enhancer (Hnisz) single-SNP annotation), analogous to the attenuated *ξ* estimates (running LDSPEC or S-LDSC^5^ using the baseline model without SNP-pair annotations produced more attenuated *τ* estimates of 5.2 *×*10^−7^ *±* 2.8 *×*10^−8^ and 5.0 *×*10^−7^ *±* 2.5 *×*10^−8^, respectively, suggesting that modeling SNP-pair annotations could partially mitigate the attenuation in these simulations; Supplementary Figure 2); estimates of total SNP-heritability and heritability enrichment were approximately unbiased, analogous to null simulations (Supplementary Figure 2b, Supplementary Table 9). Fifth, distinct from estimates of total SNP-heritability, estimates of total heritability shrinkage (total SCV divided by total SNP-heritability) were significantly smaller than 1 but attenuated towards 1 (0.80*±* 0.01, true value 0.56), consistent with the attenuation of *ξ* estimates (Supplementary Figure 2b, Supplementary Table 9). Sixth, jackknife standard errors for all quantities were well-calibrated, analogous to null simulations (Supplementary Figure 3b).

We performed 5 secondary analyses. First, we performed null and causal simulations at a lower value of SCV (0.2 instead of 0.5). Analogous to our primary simulations, LDSPEC produced approximately unbiased estimates of *ω, ξ*, and *ξ*^∗^ in null simulations, and produced significantly negative but attenuated estimates of *ω, ξ*, and *ξ*^∗^ for a subset of causal SNP-pair annotations in causal simulations (slightly biased estimates of *ω, ξ, ξ*^∗^ for other SNP-pair annotations) (Supplementary Figure 4). Second, we performed null and causal simulations at a lower value of causal SNP proportion (0.1 instead of 0.2). Analogous to our primary simulations, LDSPEC produced approximately unbiased estimates of *ξ* and *ξ*^∗^ in null simulations (though estimates of *ω* were slightly biased), and produced significantly negative but attenuated estimates of *ω, ξ*, and *ξ*^∗^ for a subset of causal SNP-pair annotations in causal simulations (Supplementary Figure 5). Third, we performed causal simulations where we specified negative *ω* values for both the 6 causal positive-LD SNP-pair annotations (as in primary simulations) and the 6 corresponding negative-LD SNP-pair annotations (vs. zero *ω* in primary simulations). Analogous to our primary causal simulations, LDSPEC produced significantly negative and slightly attenuated estimates of *ω, ξ*, and *ξ*^∗^ for a subset of causal SNP-pair annotations (with slightly biased estimates of *ω* and *ξ* for other SNP-pair annotations); the estimates were less attenuated, suggesting that LDSPEC was more effective when the positive-LD and negative-LD strata of the same SNP-pair annotation had the same *ω* (Supplementary Figure 6a). Fourth, we performed causal simulations where we specified positive *ω* values for both the 6 causal positive-LD SNP-pair annotations (vs. negative *ω* in primary simulations) and the 6 corresponding negative-LD SNP-pair annotations (vs. zero *ω* in primary simulations). Analogous to our primary causal simulations, LDSPEC produced significantly positive and slightly attenuated estimates of *ω, ξ*, and *ξ*^∗^ for a subset of causal SNP-pair annotations; once again, the estimates were less attenuated, suggesting that LDSPEC was more effective when the positive-LD and negative-LD strata of the same SNP-pair annotation had the same *ω* (Supplementary Figure 6b). Fifth, we applied LDSPEC to the primary null and causal simulation data using LD scores and directional LD scores that were computed with smaller window sizes (1Mb, 3Mb, 5Mb, instead of 10Mb). LDSPEC produced more biased estimates of *ξ*, heritability, and heritability enrichment as the window size decreased (Supplementary Figure 7).

We conclude that LDSPEC is well-calibrated in null simulations and produces attenuated estimates of nonzero SNP-pair effect correlations in causal simulations.

### Analysis of 70 diseases and complex traits

We applied LDSPEC with the baseline-SP model to publicly available summary statistics and in-sample LD of 70 diseases and complex traits (29 independent diseases/traits) from the UK Biobank^37^ (Supplementary Table 1; see Data availability), analyzing 136 SNP-pair annotations (Table 1). For each SNP-pair annotation, estimates were meta-analyzed across the 29 independent diseases/traits using random-effects meta-analysis, analogous to previous studies^5,8^ (Methods). Statistical significance was assessed via a Bonferroni p-value threshold, correcting for the number of hypotheses tested. Analysis of each UK Biobank disease/trait required roughly 12 hours for a single-core CPU, and required roughly 128GB of memory (Methods).

We first discuss results for the 3 proximity-based SNP-pair annotations (12 annotations when stratified by MAF and LD; Table 1). Results are reported in Figure 2 and Supplementary Table 14. First, for low-frequency positive-LD SNP-pair annotations, we detected strongly and significantly negative (*P*<0.05/136) SNP-pair effect correlations (*ξ*) for 0-100bp and 1-10kb SNP-pair annotations (−0.37 *±* 0.09 and −0.07 *±* 0.01; negative but non-significant estimate for 100bp-1kb). The negative *ξ* between positive-LD SNP pairs can potentially be explained by linkage masking^23^ (also see ref.^28^), whereby haplotypes containing linked SNPs with opposite effects on disease escape negative selection. Specifically, a haplotype harboring two SNPs with opposite effects on disease/trait may have a reduced aggregate effect on fitness in individuals carrying that haplotype, e.g., under stabilizing selection^38,39,46,47^. The more strongly negative *ξ* for SNP pairs at closer genomic distances may be partly because the magnitude of LD slightly decays with distance (e.g., average *r* of 0.69, 0.64, 0.55 for common positive-LD 0-100bp, 100-1kb, 1-10kb, resp., Supplementary Table 4), reducing linkage masking effects, but predominantly because nearby SNPs are more likely to have shared functional roles (e.g., median of 541bp for mean segment length across functional annotations in Supplementary Table 1 of ref.^5^); SNP pairs with similar functional roles and opposite effects on a given disease are likely to also have opposite effects on pleiotropic traits underlying pleiotropic selection^38^ (but this is less likely for SNP pairs with different functional roles). Second, for common positive-LD SNP-pair annotations, our estimate of *ξ* was negative with suggestive significance (*P*=0.001 > 0.05/136) for the 0-100bp SNP-pair annotation (−0.15 *±* 0.04; non-significant estimates for 100bp-1kb and 1-10kb). Common positive-LD SNP-pair annotations had less negative *ξ* estimates than their low-frequency counterparts (significantly positive difference for 1-10kb, *P*<0.05/68; positive but non-significant differences for the remaining 2 comparisons; Supplementary Table 15), perhaps because common SNPs have smaller per-allele effects on disease and fitness than low-frequency SNPs^11,13,16^, limiting the impact of linkage masking. Third, common and low-frequency negative-LD SNP-pair annotations had less negative *ξ* estimates than their positive-LD counterparts (significantly positive differences for common 0-100bp, *P*<0.05/68; positive but non-significant differences for the remaining 5 comparisons; Supplementary Table 15), consistent with linkage masking, which implicates a negative SNP-pair effect correlation for positive-LD SNP pairs and a less negative or weakly positive SNP-pair effect correlation for negative-LD SNP pairs (see *Forward simulations recapitulate empirical findings*.)

**Figure 2.**
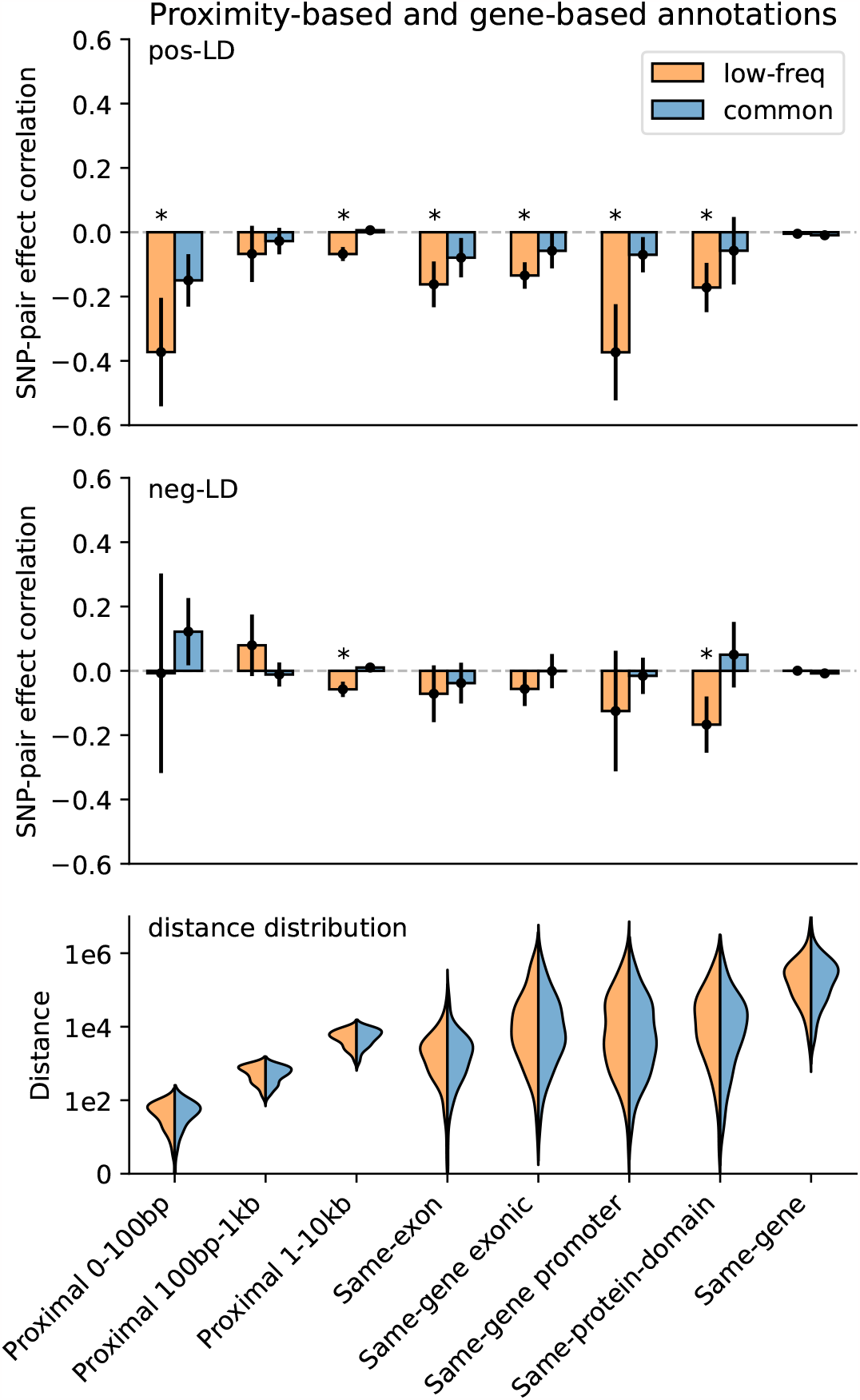
Estimates of SNP-pair effect correlation (*ξ*) across 29 independent diseases and complex traits for proximity-based and gene-based SNP-pair annotations. We report meta-analyzed *ξ* estimates across 29 independent diseases for 3 proximity-based and 5 gene-based SNP-pair annotations. Results are shown for the low-frequency positive-LD, common positive-LD, low-frequency negative-LD, and common negative-LD SNP-pair annotations, respectively (upper and middle panels). Error bars denote 95% confidence intervals. “*” denotes statistical significance after multiple testing correction (*P*<0.05/136). The lower panel shows the distance distribution across SNP pairs for each annotation, where positive-LD and negative-LD SNP pairs are combined because their distributions are similar. The large distance for the same-gene promoter SNP-pair annotation is because a gene may have multiple promoter regions due to alternative splicing^70^. Numerical results are reported in Supplementary Table 14.

We next discuss results for the 5 gene-based SNP-pair annotations (20 annotations when stratified by MAF and LD; Table 1). Results are reported in Figure 2 and Supplementary Table 14. First, for low-frequency positive-LD SNP-pair annotations, we detected strongly and significantly negative (*P*<0.05/136) SNP-pair effect correlations (*ξ*) for same-exon, same-gene exonic, same-gene promoter, and same-protein-domain SNP-pair annotations (−0.16 *±* 0.04, −0.13 *±* 0.02, −0.37*±* 0.08, and −0.17 *±* 0.04; estimates of excess SNP-pair effect correlation (*ξ*^∗^) were very similar to estimates of *ξ* for these SNP-pair annotations due to their large genomic distances (implying a close to zero expected value of *ξ* for distance-matched SNP pairs) (Supplementary Table 13). The strongly negative *ξ* (and *ξ*^∗^) estimates are consistent with shared functional roles for SNP pairs in these gene-based annotations; the same-gene promoter SNP-pair annotation had the most negative *ξ* estimate, perhaps because promoter SNPs can either increase or decrease gene expression levels^48^, supporting masking effects on gene expression, disease/trait, and fitness. Second, for common positive-LD SNP-pair annotations, *ξ* estimates were non-significant and less negative than their low-frequency counterparts (significantly positive difference for same-gene promoter, *P*<0.05/68; positive but non-significant differences for 4 of 5 comparisons; Supplementary Table 15), analogous to results for proximity-based SNP-pair annotations. Third, common and low-frequency negative-LD SNP-pair annotations had less negative *ξ* estimates than their positive-LD counterparts (significantly positive differences for 9 of 10 comparisons, *P*<0.05/68; positive but non-significant difference for the remaining 1 comparison; Supplementary Table 15), analogous to results for proximity-based SNP-pair annotations.

Finally, we discuss results for the 7 functional 0-100bp and 19 functional 0-1kb SNP-pair annotations (e.g., pairs of H3K27ac SNPs with distance 0-100bp; 104 annotations when stratified by MAF and LD; Table 1). We primarily focus on excess SNP-pair effect correlations (*ξ*^∗^) to assess information specific to these functional annotations. *ξ*^∗^ estimates are reported in Figure 3 and Supplementary Table 16; corresponding *ξ* estimates are reported in Supplementary Figure 8 and Supplementary Table 13. First, for low-frequency positive-LD SNP-pair annotations, we detected strongly and significantly negative (*P*<0.05/136) *ξ*^∗^ for 9 of 19 functional 0-1kb SNP-pair annotations (e.g., −0.24 *±* 0.02 for H3K27ac 0-1kb; significantly positive for Repressed 0-1kb, 0.21 *±* 0.10, *P*<0.05/136; non-significant for the remaining 9 functional 0-1kb and all 7 functional 0-100bp). SNP pairs in these SNP-pair annotations have stronger effects on disease^5^ and are likely to have similar functional roles, thus are expected to be more strongly impacted by linkage masking (exception: the significantly positive *ξ*^∗^ estimate for the low-frequency positive-LD Repressed 0-1kb SNP-pair annotation (corresponding *ξ* estimate non-significant) is likely because SNP pairs in this annotation have weaker effects on disease^5^ and are likely to have weaker effects on fitness, thus expected to be less strongly impacted by linkage masking). Interestingly, low-frequency positive-LD functional 0-100bp SNP-pair annotations had less negative *ξ*^∗^ estimates than the corresponding functional 0-1kb SNP-pair annotations (significantly positive differences for H3K27ac and Transcribed, *P*<0.05/7; non-significant for the remaining 5; Supplementary Table 17); SNP pairs at very short genomic distances may generally have shared functional roles supporting linkage masking regardless of functional annotation, limiting the difference in *ξ* between functional SNP pairs and other distance-matched SNP pairs. Second, for common positive-LD SNP-pair annotations, we detected significantly negative (*P*<0.05/136) *ξ*^∗^ for only 4 of 19 functional 0-1kb SNP-pair annotations (e.g., −0.05 *±* 0.01 for H3K27ac 0-1kb; non-significant for the remaining 15 functional 0-1kb and all 7 functional 0-100bp). Common positive-LD functional SNP-pair annotations had less negative *ξ*^∗^ estimates than their low-frequency counterparts (significantly positive differences for 12 out of 26 comparisons, *P*<0.05/68; Supplementary Table 17), analogous to results for proximity-based SNP-pair annotations. Third, common and low-frequency negative-LD functional SNP-pair annotations had less negative *ξ*^∗^ estimates than their positive-LD counterparts (significantly positive differences for 36 of 38 functional 0-1kb (and 0 of 14 functional 0-100bp), significantly negative difference for common Repressed 0-1kb, *P*<0.05/68; Supplementary Table 17), analogous to results for proximity-based SNP-pair annotations; 5 of 19 common negative-LD functional 0-1kb SNP-pair annotations had weakly but significantly positive (*P*<0.05/136) *ξ* estimates (Supplementary Figure 8), perhaps because SNP pairs with concordant effects are more likely to be on different haplotypes to have a smaller aggregate impact on fitness under stabilizing selection.

**Figure 3.**
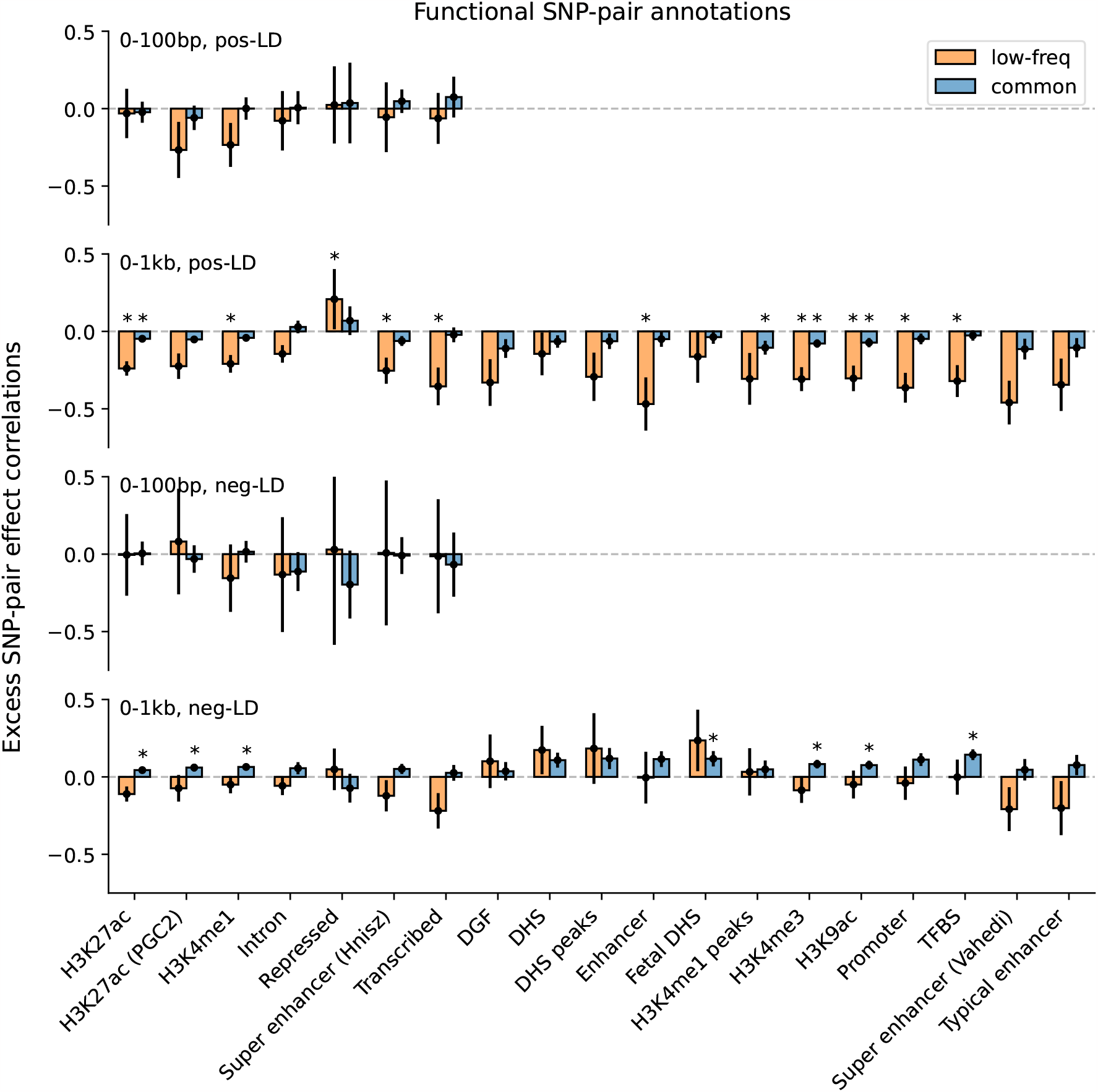
Estimates of excess SNP-pair effect correlation (*ξ*^∗^) across 29 independent diseases and complex traits for functional SNP-pair annotations. We report meta-analyzed *ξ*^∗^ estimates across 29 independent diseases for 7 functional 0-100bp and 19 functional 0-1kb SNP-pair annotations. Results are shown for the positive-LD 0-100bp, positive-LD 0-1kb, negative-LD 0-100bp, and negative-LD 0-1kb SNP-pair annotations in the 4 panels, respectively, and are stratified by MAF in each panel. Error bars denote 95% confidence intervals. “*” denotes statistical significance after multiple testing correction (*P*<0.05/136). Numerical results are reported in Supplementary Table 16.

We investigated whether excess SNP-pair effect correlations (*ξ*^∗^) were larger for functional SNP-pair annotations with larger disease heritability enrichments for the underlying functional single-SNP annotations; we hypothesized that this might be the case, because pairs of SNPs with more strongly enriched heritability and shared functional roles are expected to be more strongly impacted by linkage masking. Results are reported in Figure 4, Supplementary Figure 9, and Supplementary Table 18. For positive-LD functional SNP-pair annotations, we observed significantly more negative (*P*<0.05/4) *ξ*^∗^ estimates for functional annotations with higher disease heritability enrichments, with a stronger effect for low-frequency SNP-pair annotations (e.g., regression slope of −0.179 *±* 0.031 for low-frequency positive-LD 0-1kb vs. −0.024 *±* 0.009 for common positive-LD 0-1kb). For negative-LD functional SNP-pair annotations, we observed significantly more positive (*P*<0.05/4) *ξ*^∗^ estimates for functional annotations with higher disease heritability enrichments (e.g., regression slope of 0.036 *±* 0.010 for common negative-LD 0-1kb; non-significant slope of -0.041 *±* 0.029 for low-frequency negative-LD 0-1kb). These results support our hypothesis that functional annotations that are more enriched for disease heritability are more impacted by linkage masking.

**Figure 4.**
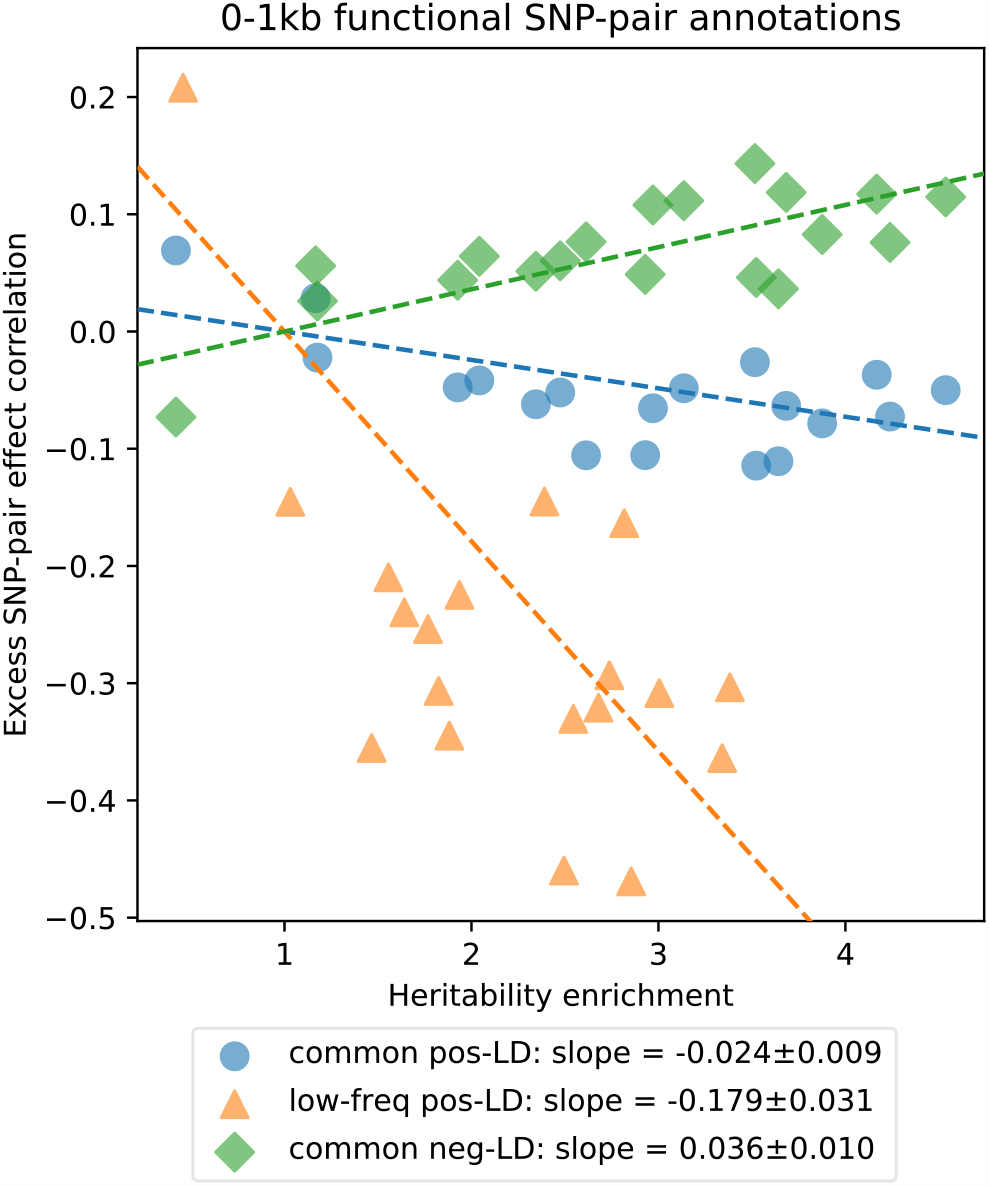
Comparison between estimates of heritability enrichment and estimates of excess SNP-pair effect correlation (*ξ*^∗^) across 19 functional 0-1kb SNP-pair annotations. Each dot represents a SNP-pair annotation, x-axis represents the meta-analyzed estimate of heritability enrichment, and y-axis represents the meta-analyzed estimate of *ξ*^∗^ (across 29 independent diseases/traits). Results are shown for the common positive-LD, low-frequency positive-LD, and common negative-LD SNP-pair annotations separately (significantly nonzero slope with *P*<0.05/4); results were not significant for the low-frequency negative-LD SNP-pair annotation (*P*>0.05/4; not shown). Regression slopes are provided with SEs in the figure legend. Complete results are reported in Supplementary Figure 9. Numerical results are reported in Supplementary Table 18.

Although most of our results reflect a meta-analysis across diseases/traits, an assessment of results for individual diseases/traits is also important. For individual diseases/traits, we detected 12 significantly nonzero (*P*<0.05/136) SNP-pair effect correlations (*ξ*), spanning 10 diseases/traits and 9 SNP-pair annotations (Supplementary Table 11); this suggests that LDSPEC can detect nonzero *ξ* for individual diseases/traits, but has limited power to do so. These findings included a significantly negative *ξ* estimate of the common positive-LD H3K4me3 0-1kb SNP-pair annotation for Monocyte Count (−0.19 *±* 0.05) and a significantly positive *ξ* estimate of the common negative-LD 0-1kb H3K4me1 SNP-pair annotation for Forced Vital Capacity (0.21*±*0.05). We assessed the heterogeneity of *ξ* estimates across 29 independent diseases/traits by computing a statistic quantifying relative excess cross-trait variance as compared to within-trait variance (Methods). Results are reported in Supplementary Table 19. The medium relative excess cross-trait variance was 4.0% across all 136 SNP-pair annotations (17.3% when restricting to the 12 proximity-based SNP-pair annotations), implying a low level of heterogeneity. We detected significant heterogeneity (*P*<0.05/136) for 1 SNP-pair annotation, the low-frequency positive-LD Repressed 0-1kb SNP-pair annotation (*P*=3.5 *×*10^−4^).

We compared LDSPEC results obtained using the baseline-SP model to results obtained using other heritability models,including the baseline-SP-proximity model (165 single-SNP annotations + 12 proximity-based SNP-pair annotations only), the baseline-SP-gene model (165 single-SNP annotations + 20 gene-based SNP-pair annotations only), and the baseline-SP-functional model (165 single-SNP annotations + 104 functional SNP-pair annotations only). We determined that each of these models produced similar *ξ* estimates as the baseline-SP model for SNP-pair annotations shared between the models (correlation of 0.96 across 136 SNP-pair correlations; non-significant difference (*P*>0.05/136) for all 136 comparisons; Supplementary Figure 10).

We conclude that positive-LD SNP pairs tend to have strongly negative SNP-pair effect correlations of disease effects, negative-LD SNP pairs tend to have less negative or weakly positive SNP-pair effect correlations, low-frequency SNP pairs tend to have stronger SNP-pair effect correlations than common SNP pairs, and SNP pairs in shared functional annotations tend to have much stronger SNP-pair effect correlations.

### Impact of SNP-pair effect correlations on SNP-heritability

We assessed the impact of SNP-pair effect correlation on SNP-heritability by estimating and comparing two closely related quantities: SNP-heritability and sum of causal effect size variances (SCV) (Methods); the two quantities may be different when causal effects are not independent (as assumed in previous work^1–21^). SNP-heritability quantifies the aggregate impact of SNPs on disease and may be more relevant to applications such as polygenic risk scores (PRS)^49,50^, whereas SCV pertains to the impact of individual SNPs on disease and may be more relevant to applications such as fine-mapping^51^.

Results are reported in Figure 5 and Supplementary Table 20. First, SNP-heritability was substantially smaller than SCV, with a regression slope of 0.89 *±* 0.01; accordingly, *heritability shrinkage*, defined as the ratio between SNP-heritability and SCV, was equal to 0.87*±* 0.02 (average across 29 independent diseases/traits). This implies that the phenomenon of negative SNP-pair effect correlations for positive-LD SNP pairs (and less negative or weakly positive SNP-pair effect correlations for negative-LD SNP pairs) can substantially impact SNP-heritability. Second, average heritability shrinkage was even stronger for certain functional annotations, e.g., 0.79*±* 0.01 for common Super enhancer (Hnisz) SNPs; average of 0.83 *±* 0.01 across the 6 common functional annotations that had enriched heritability (heritability enrichment >1) and were large enough to be included in both 0-100bp and 0-1kb SNP-pair annotations (implying more accurate modeling of heritability shrinkage) and 0.84 *±* 0.01 across the corresponding 6 low-frequency functional annotations.

**Figure 5.**
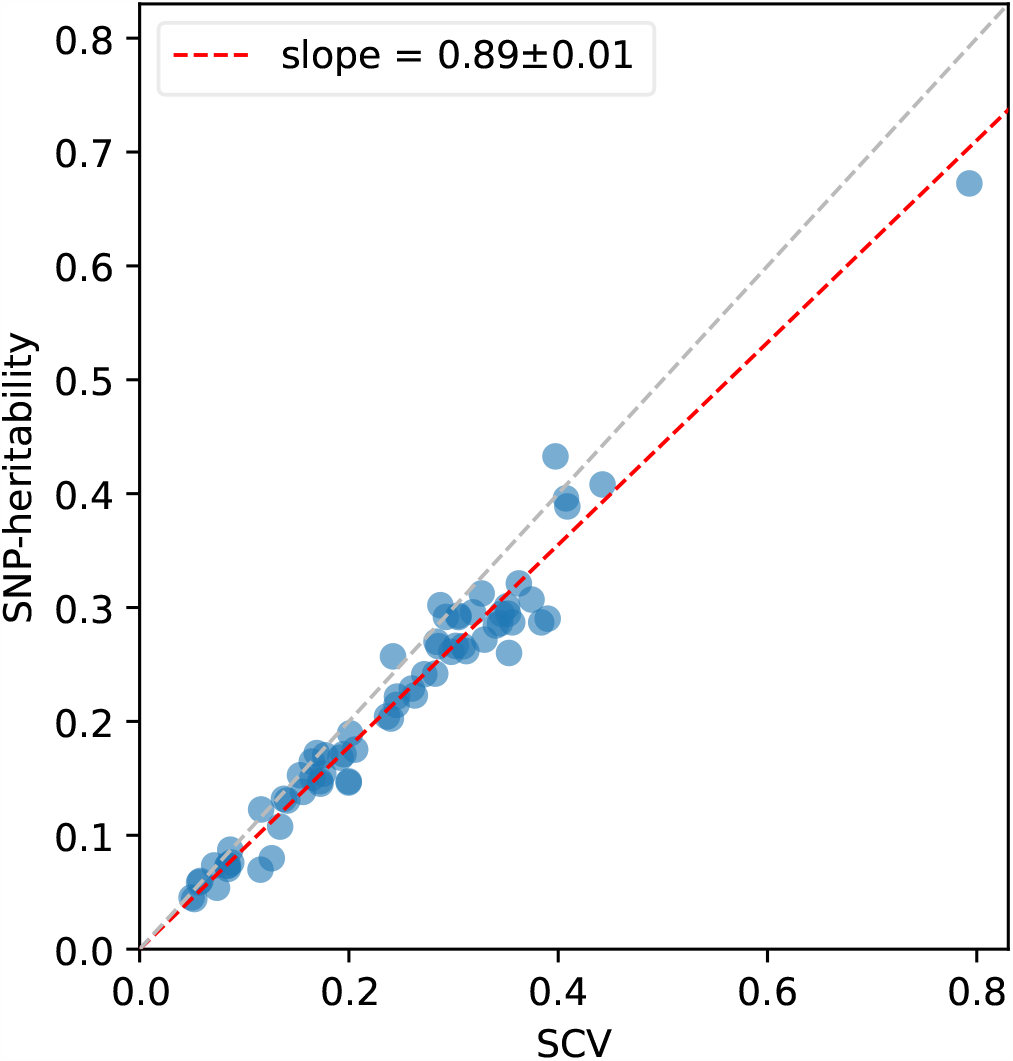
Comparison between estimates of SCV and estimates of SNP-heritability across 70 diseases and complex traits. Each dot represents a disease/trait, x-axis represents the estimate of SCV, and y-axis represents the estimate of SNP-heritability. Regression slope was obtained by linear regression without intercept across 29 independent diseases/traits. Numerical results are reported in Supplementary Table 20.

We performed 3 secondary analyses. First, we assessed the impact of modeling SNP-pair effect correlations on genome-wide SNP-heritability estimates; we determined that modeling SNP-pair effect correlations had a limited impact, as models that do not account for SNP-pair effect correlations produced similar estimates (Supplementary Figure 11a). Second, we assessed the impact of modeling SNP-pair effect correlations on estimates of heritability enrichment for single-SNP annotations; again, we determined that modeling SNP-pair effect correlations had a limited impact, as models that do not account for SNP-pair effect correlations produced similar estimates (Supplementary Figure 11b). Third, we confirmed that LDSPEC and S-LDSC^5,8^ (using the baseline model without SNP-pair annotations) produced similar estimates of each single-SNP annotation’s contribution to per-SNP heritability (*τ*), as well as genome-wide SNP-heritability (Supplementary Figure 11c,d).

We conclude that SNP-heritability is systematically smaller than SCV across diseases/traits, and that this heritability shrinkage is stronger for functionally important annotations.

### Forward simulations under stabilizing selection recapitulate empirical findings

Our finding that positive-LD SNP pairs tend to have negative SNP-pair effect correlations can potentially be explained by linkage masking, whereby haplotypes containing linked SNPs with opposite effects on disease have reduced effects on fitness and escape negative selection^23,28^. To test this hypothesis, we performed forward simulations of a quantitative trait under stabilizing selection, in which alleles that either increase or decrease the value of the phenotype are selected against^38,39^. In our primary simulations, we assumed a constant population size with 10,000 diploid individuals, mutation rate *μ* = 1 *×*10^−8^, and fitness function (defined as the relationship between fitness and trait effect size of an allele) consistent with strong stabilizing selection (width of fitness function = 2; Supplementary Figure 12a); other settings were also evaluated. We assessed the (true) SNP-pair effect correlations (*ξ*) of SNP-pair annotations stratified by MAF and LD at different distances. Further details of the forward simulation framework are provided in the Methods section.

Results are reported in Figure 6 and Supplementary Table 21. We determined that positive-LD 0-100bp, 100bp-1kb, and 1-10kb SNP pairs had substantially negative SNP-pair effect correlations whereas negative-LD 0-100bp, 100bp-1kb, and 1-10kb SNP pairs had weakly positive SNP-pair effect correlations, which is consistent with linkage masking and qualitatively consistent with results for real diseases/traits (Figure 2). We did not observe a sharp decay of *ξ* with distance as in real data (Figure 2), perhaps because we did not simulate more proximal SNPs to have shared functional roles, which is the case in real data (Supplementary Table 1 of ref.^5^). Under stabilizing selection, SNP pairs with discordant effects on the trait (for derived alleles) will have strongly positive LD, because haplotypes containing both derived alleles or both ancestral alleles are less susceptible to selection (than haplotypes containing one derived allele and one ancestral allele). On the other hand, SNP pairs with concordant effects on the trait (for derived alleles) will have weakly negative LD, because haplotypes containing both derived alleles are more susceptible to selection but haplotypes containing both ancestral alleles are less susceptible to selection (than haplotypes containing one derived allele and one ancestral allele). These consequences are consistent with the “Bulmer effect”, in which stabilizing selection reduces the phenotypic variance in each generation by weeding out extreme deviations from the norm^46,47^.

**Figure 6.**
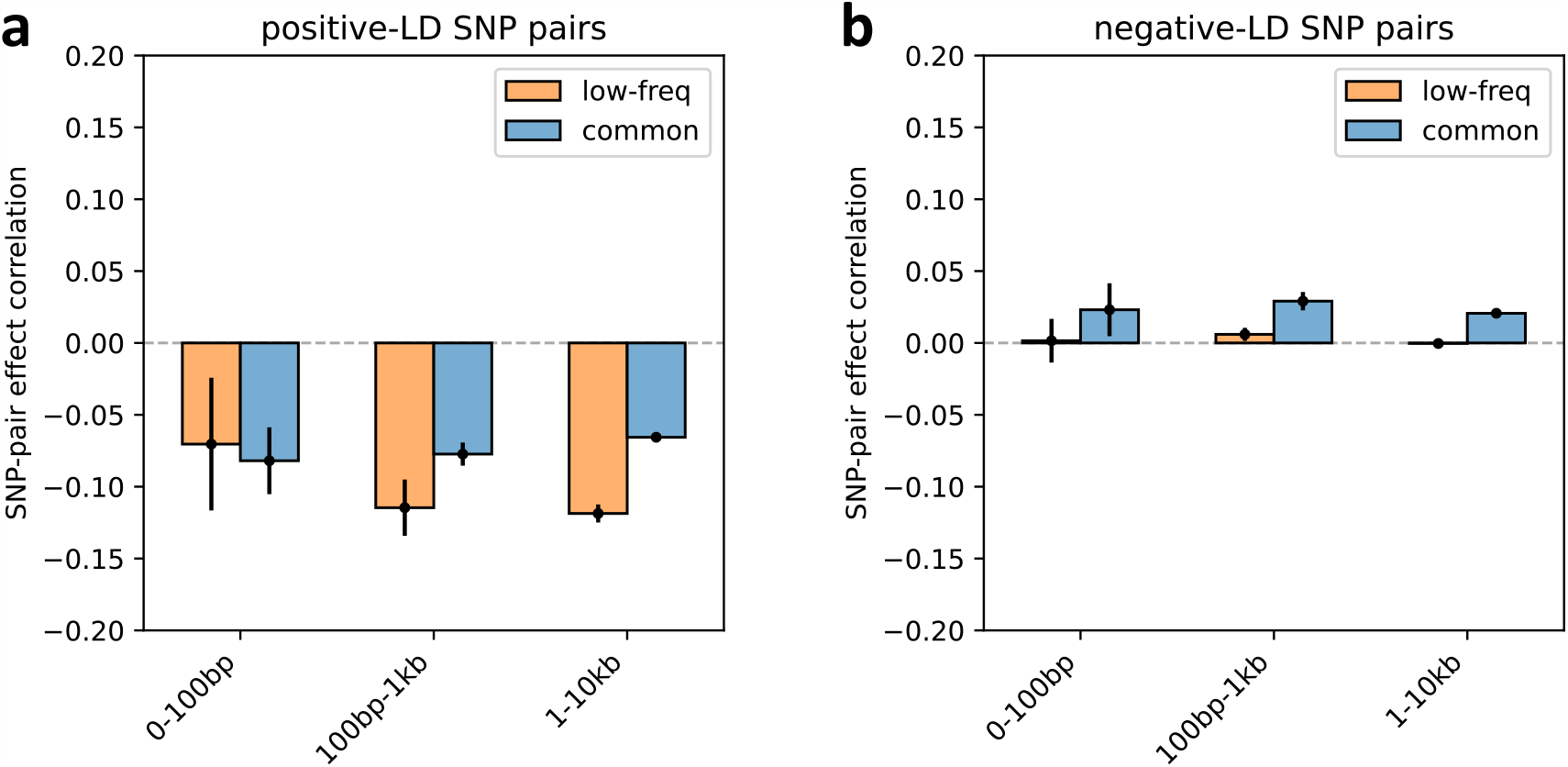
SNP-pair effect correlation (*ξ*) in forward evolutionary simulations with stabilizing selection. Panels a and b report values of *ξ* for positive-LD and negative-LD SNP pairs, respectively. For each panel, results are reported for common and low-frequency SNP pairs separately, stratified into 0-100bp, 100bp-10kb, and 1-10kb distance bins. Error bars denote 95% CIs. Numerical results are reported in Supplementary Table 21.

Accordingly, we determined that SNP pairs with opposite trait effects (for derived alleles) tended to be in strongly positive LD, and SNP pairs with concordant trait effects (for derived alleles) tended to be in weakly negative LD (Supplementary Figure 12b). The level of LD was relatively low when the disease/trait effects were either very small or very large, perhaps because small-effect SNPs are less impacted by stabilizing selection, and large-effect SNPs are efficiently removed from the population before the emergence of a second SNP masking the first SNP’s trait effect. LD was not significantly different from zero for neutral SNP pairs with at least one zero-effect SNP (Supplementary Figure 12b), consistent with the hypothesis that negative *ξ* arises only under selection. We also performed simulations with other selection strengths (width of the fitness function: 4 for moderate selection and 1 *×*10^6^ for no selection, instead of 2 for strong selection in primary simulation). Results were similar for moderate selection vs. strong selection, but the LD between SNP pairs with correlated effects disappeared under no selection, consistent with our expectation (Supplementary Figure 12b).

In summary, our results suggest that a model of stabilizing selection on a complex trait can potentially explain the patterns we observe in real data, providing an evolutionary explanation for our findings.

## Discussion

We have developed LDSPEC, a method that analyzes summary statistics and in-sample LD to estimate correlations of causal disease effect sizes for pairs of nearby SNPs, depending on their functional annotations. We recommend applying LDSPEC using the baseline-SP model, which contains 165 single-SNP annotations^11^ and 136 new SNP-pair annotations, including 12 proximity-based, 20 gene-based, and 104 functional SNP-pair annotations. We have shown that LDSPEC is approximately unbiased and well-calibrated in null simulations and capable of detecting nonzero SNP-pair effect correlations (with attenuated estimates) in causal simulations. Applying LDSPEC with the baseline-SP model to 70 UK Biobank diseases and complex traits^37^, we detected strongly and significantly nonzero SNP-pair effect correlations for nearby SNP pairs that decayed with distance. We determined that positive-LD SNP pairs had strongly negative disease-effect correlations, that negative-LD SNP pairs had less negative or weakly positive disease-effect correlations, and that SNP pairs in shared functional annotations that were enriched for disease heritability had stronger disease-effect correlations that spanned longer distances. As a consequence, SNP-heritability is systematically smaller than the sum of causal effect size variances, particularly for certain functional annotations. The negative SNP-pair effect correlations between positive-LD SNP pairs can potentially be explained by linkage masking, whereby haplotypes containing linked SNPs with opposite effects on disease have a reduced aggregate effect on fitness and escape negative selection. Forward simulations showed that our findings are consistent with an evolutionary model involving stabilizing selection.

To our knowledge, no published study has systematically investigated SNP-pair effect correlations in genome-wide data. Our work expands upon an unpublished preprint^40^, which contained key ideas and derivations and detected SNP-pair effect correlations for extremely-short-range SNP pairs (0-100bp) that varied with LD. We note 4 important differences between our work and ref.^40^. First, our work stratifies SNP pairs by MAF and functional annotations. Second, our work identifies SNP-pair effect correlations at larger genomic distances (up to tens of kilobases). Third, our work performs evolutionary forward simulations to interpret our findings. Fourth, our work introduces improved methodology: LDSPEC uses a more accurate model^15^ for per-SNP heritability (165 baseline-LF single-SNP annotations^11^ vs. 26 MAF-and-LD single-SNP annotations in ref.^40^); LDSPEC adopts a principled estimator for SNP-pair effect correlations, whereas ref.^40^ uses a two-step heuristic assuming per-SNP heritability to be the same across SNPs; and LDSPEC more accurately computes LD scores and directional LD scores using a much larger LD window (10Mb vs. 1Mb) (leveraging an efficient implementation).

Our findings have several implications for future work. First, our findings challenge the widespread assumption of independent causal SNP-to-disease effects in studies of disease and complex trait architectures^1–21^. We have shown that modeling SNP-pair effect correlations distinguishes total SNP-heritability from the sum of causal SNP-to-trait effect size variances. Despite the limited impact of modeling SNP-pair effect correlations on estimates of SNP-heritability and heritability enrichment, its impact on other genetic architecture parameters (e.g., parameters related to polygenicity^13,14,17,19^ or selection^11,13,16,20^) remains to be assessed. Second, our findings motivate further prioritization of joint association testing methods that increase statistical power in the presence of linkage masking^23,28,52^. Third, our findings motivate the development of improved fine-mapping methods to disentangle linkage-masked SNPs by modeling SNP-pair effect correlations; incorporating functional annotations^43,51–54^ (including SNP-pair annotations) and analyzing data from diverse populations with different LD patterns^55–57^ will likely remain valuable. Fourth, negative SNP-pair effect correlations may contribute to poor cross-population transferability of polygenic risk scores (PRS)^49,50,58–60^, as linked SNPs with opposite effects in one population may not be linkage-masked in a different population due to different LD patterns. Ongoing efforts to improve cross-population PRS^61,62^ may benefit from modeling SNP-pair effect correlations.

We note several limitations of our work. First, LDSPEC produces attenuated estimates of SNP-pair effect correlations in causal simulations, possibly because there is a high level of collinearity of directional LD scores between SNP-pair annotations, and it is challenging to distinguish *ξ* between SNP-pair annotations with highly correlated directional LD scores; however, LDSPEC is unbiased and well-calibrated in null simulations. Second, LDSPEC attains incomplete power in some settings, including simulations (Figure 1b) and analyses of individual diseases/traits (Supplementary Tables 10,11); an important future direction is to improve the power of LDSPEC, e.g., by incorporating products of z-scores of nearby SNPs. Third, we only considered binary SNP-pair annotations in this work; an important future direction is to extend LDSPEC to incorporate continuous SNP-pair annotations, analogous to incorporation of continuous single-SNP annotations in S-LDSC^8^. Fourth, although we have shown via forward simulations that stabilizing selection can produce the negative SNP-pair effect correlations observed in real data, we currently cannot exclude the possibility that this could be produced by other evolutionary mechanisms. For example, Hill–Robertson interference^8,22^ can create negative LD for pairs of deleterious SNPs (concordant effects on fitness) and antagonistic epistasis can create positive LD between SNP pairs^26^. Stabilizing selection may be a more plausible explanation, because Hill–Robertson interference is less relevant to SNP pairs with opposite effects and the impact of epistatis on disease is hypothesized to be small^63–65^. Nonetheless, investigating the impact of a broad set of evolutionary models on SNP-pair effect correlations is an important future direction. Fifth, we have estimated SNP-pair effect correlations for low-frequency and common variants, but not for rare variants (for which LDSPEC is underpowered due to a lower level of LD between rare SNP pairs). Investigating SNP-pair effect correlations for rare variants (which have often been reported to have concordant effects^29–34^, motivating the development of rare variant burden tests^52,66,67^) is an important future direction. Sixth, analogous to other studies that employ linear complex trait models^1–21^, we have not investigated the potential impact of epistatic interactions on our estimates; however, the impact of epistatic interaction on these models is hypothesized to be small^63–65^. Seventh, we have not assessed the impact of unmodeled causal variants that are missing from the data on our estimates. However, shared tagging of unmodeled causal variants could produce spurious positive effect correlations between positive-LD SNP pairs, but would not be expected to produce the negative effect correlations that we report here. Eighth, we have analyzed “in.white.British.ancestry.subset” samples from the UK Biobank, but an important future direction is to extend our analyses to cohorts of diverse genetic ancestry^68,69^. Despite these limitations, our work provides a comprehensive genome-wide assessment of SNP-pair effect correlations of causal disease effect sizes across MAF, LD, and functional annotations.

## Acknowledgements

We are grateful to Kangcheng Hou, Kushal Dey, Ali Akbari, Luke O’Connor, Armin Schoech, Corbin Quick, Xihao Li, Hui Li, Tiffany Amariuta, Karthik Jagadeesh, Katherine Siewert-Rocks, Jordan Rossen, Elizabeth Dorans, Xihong Lin, and Soumya Raychaudhuri for their helpful discussions. This research was conducted using the UK Biobank resource under application no. 16549 and was funded by National Institutes of Health (NIH) grants U01 HG009379, R01 MH101244, R37 MH107649, U01 HG012009 and R01 HG006399. C.C. was funded by the NIGMS 5T32GM007748-44. The funders had no role in study design, data collection and analysis, decision to publish or preparation of the manuscript.

## Author Contributions Statement

M.Z. and A.P. designed the study and developed statistical methodologies. M.Z. analyzed the genetic data with assistance from A.D., C.C. A.D. and C.C. performed forward evolutionary simulations. E.K., A.S., B.S., P.L., S.G., and S.S. provided expert guidance and feedback on analysis, results and biological interpretations. M.Z., A.D., C.C., S.S., and A.P. wrote the manuscript with feedback from all authors.

## Competing Interests Statement

The authors declare no competing interests.

## Methods

### Modelling SNP-pair effect correlations

We considered *N* individuals, *M* SNPs, and assume a polygenic model^1,71^

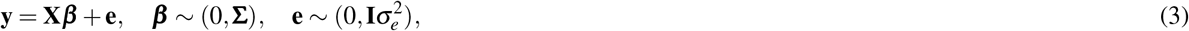

where **y** ∈ ℝ^*N*^ is a quantitative phenotype, **X**∈ ℝ^*N×M*^ is the standardized genotype, ***β*** ∈ ℝ^*M*^ is the SNP causal effects on phenotype, and **e** ∈ ℝ^*N*^ is the environmental factor. We model **X** as fixed and model ***β*** and **e** as random variables independent of each other. Previous work has assumed independent SNP-to-phenotype effects^1–21^ (implying elements of ***β*** are independent), but our model allows SNP-to-phenotype effects to be correlated by assuming a general covariance ***β*** ∼ (0, **Σ**). We standardize^71^ **X** as 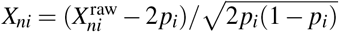, where 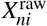 is the number of derived alleles for individual *n* and SNP *i*, and *p*_*i*_ is the derived allele frequency of SNP *i*.

We consider *C* binary/continuous single-SNP annotations, where *a*_*c*_(*i*) ∈ ℝrepresents the value of annotation *c* for SNP *i*. We consider *K* binary SNP-pair annotations, where *G*_*k*_(*i, j*) ∈ {0, 1} indicates if SNP pair (*i, j*) is in the annotation (we set diagonal elements *G*_*k*_(*i, i*) = 0 for modelling convenience). We model the SNP causal effect covariance as a linear combination of contributions from single-SNP annotations and SNP-pair annotations:

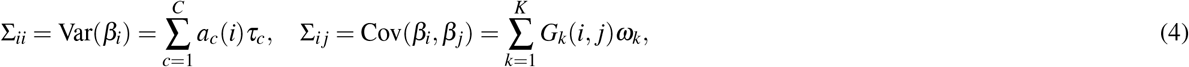

where *τ*_*c*_ represents the contribution of single-SNP annotation *c* to per-SNP heritability, and *ω*_*k*_ represents the contribution of SNP-pair annotation *k* to per-SNP-pair covariance. Analyzing standardized effect sizes (as in this paper) may produce slightly different results compared to analyzing non-standardized (per-allele) effect sizes, as the two analyses, together with model (4), imply different MAF-dependent genetic architectures.

### Inference via LDSPEC

Let 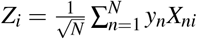 be the summary association statistic for SNP *i* and 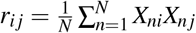 be the signed in-sample LD between SNPs *i* and *j*. Then the chi-square statistic 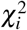 is equal to 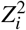. Under the correlated SNP effect model (Equations (3),(4)),

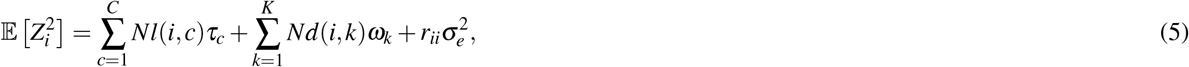

where 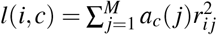 is the LD score of SNP *i* for single-SNP annotation *c* and 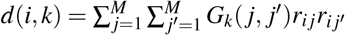 is the directional LD score of SNP *i* for SNP-pair annotation *k*. Please see the Supplementary Note for more details.

We use all SNPs in the data set as both reference SNPs (for computing LD and directional LD scores) and regression SNPs (for estimating *τ*_*c*_ and *ω*_*k*_ via regression). We prefer in-sample LD over external LD reference panels because external LD data sets may have smaller sample sizes and may not match the GWAS cohort, potentially reducing power and introducing estimation bias. For computational tractability, we approximate the LD and directional LD scores using SNPs in an adjacent 10Mb window; using a smaller window may introduce estimation biases (Supplementary Figure 7). We use two sets of regression weights similar to previous work^4^: LD score weights proportional to 1*/l*(*i*) accounting for dependency between regression SNPs and heteroskedasticity weights proportional to 1*/*(*Nl*(*i*)*/M* + 1)^2^ (approximating 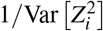), where 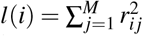 is the LD score of SNP *i* and is estimated using reference SNPs in the adjacent 10Mb window. We estimate the covariance of estimates of *τ*_*c*_ and *ω*_*k*_ using a genomic block jackknife with 100 equally-sized blocks of adjacent SNPs; estimates of *τ*_*c*_ and *ω*_*k*_ are approximately normally distributed.

LDSPEC further estimates a number of parameters for single-SNP annotations and SNP-pair annotations. Let *a*_*c*_ = *i* : *a*_*c*_(*i*) = 1} be the set of SNPs in a binary single-SNP annotation *c* and *G*_*k*_ = (*i, j*) : *G*_*k*_(*i, j*) = 1} be the set of SNP pairs in a SNP-pair annotation *k*.

1. Heritability of a single-SNP annotation 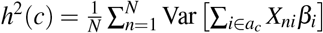. It holds that 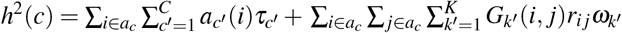 (second term is 0 when SNP effects are independent; see Supplementary Note for more details). For computational efficiency, we approximate the coefficient of *ω*_*k*_*′* in the second term as 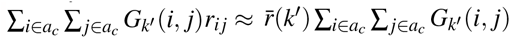, where 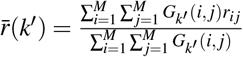 is the average signed LD across SNP pairs in *G*_*k*_*′* and can be precomputed (see Data availability).
2. Sum of causal effect size variance (SCV) of a single-SNP annotation 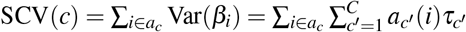 is equal to *h*^2^(*c*) when SNP effects are independent.
3. Heritability enrichment of a single-SNP annotation *c*^11^. For a common single-SNP annotation *c*, the common heritability enrichment is 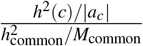, where 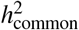 is the common SNP heritability and *M* is the number of common SNPs. We define and estimate low-frequency heritability enrichment for a low-frequency single-SNP annotation similarly.
4. Heritability shrinkage of a single-SNP annotation 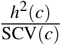.
5. Total SNP-pair effect covariance of a SNP-pair annotation 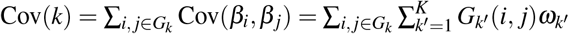.
6. SNP-pair effect correlation of a SNP-pair annotation 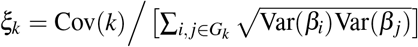, where 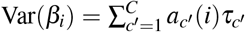.
7. Total excess SNP-pair effect covariance of a SNP-pair annotation 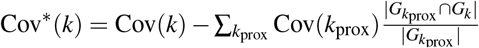, where, for a heritability model with non-overlapping proximity-based SNP-pair annotations (such as baseline-SP), ∑_*k*prox_ sums over the non-overlapping proximity-based SNP-pair annotations. Cov^∗^(*k*) = 0 for proximity-based annotations by definition.
8. Excess SNP-pair effect correlation of a SNP-pair annotation 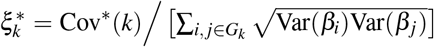, where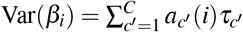.

Heritability, SCV, total SNP-pair effect covariance, and excess total SNP-pair effect covariance are linear in *τ*_*c*_*′* and *ω*_*k*_*′* (therefore approximately normal); we estimate their SE and further compute z-scores to test for significance using the covariance of estimates of *τ*_*c*_*′* and *ω*_*k*_*′*. Since heritability enrichment may not be normally distributed, analogous to previous work, we test for significant enrichment (≠1) by testing whether _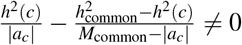_, which is linear in *τ*_*c*_*′* and *ω*_*k*_*′* (therefore approximately normal). Since heritability shrinkage may not be normally distributed, we test for significant shrinkage ≠1 by testing whether *h*^2^(*c*) − SCV(*c*) ≠0, which is linear in *τ*_*c*_*′* and *ω*_*k*_*′* (therefore approximately normal). Since *ξ*_*k*_ (resp. 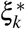) may not be normally distributed, we test for significantly nonzero *ξ*_*k*_ (resp. 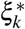) using the p-value for nonzero Cov(*k*) (resp. Cov^∗^(*k*)). We also report jackknife SE for heritability enrichment, heritability shrinkage, *ξ*_*k*_, and 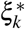, even though this is not what we use to assess significance.

The computational cost for LDSPEC to analyze one UK Biobank disease/trait (14,820,648 SNPs) was roughly 12 hours for a single-core CPU, and roughly 128GB of memory; this assumes precomputed LD and directional LD scores (which need to be computed only once for all diseases/traits analyzed).

### Genotype data

We considered 337,426 unrelated “in.white.British.ancestry.subset” individuals and 70 diseases and complex traits from the UK Biobank^37^ (average *N*=305,646, z-score >5 for nonzero SNP-heritability; Supplementary Table 1). The subset of 29 independent diseases/traits (average *N*=298,430) was selected to have pairwise genetic correlation^7^ 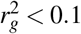. We considered the set of 14,820,648 UK Biobank imputed SNPs (version “imp_v3” from ref.^37^) with MAF ≥ 0.1% and INFO score ≥ 0.6, similar to previous work^11,43^. This set of SNPs was used as both the regression SNPs and reference SNPs in the LDSPEC analysis. We considered disease effects defined with respect to derived alleles of SNPs. To determine the ancestral allele (opposite of the derived allele) at each variant site, we obtained a whole genome alignment of the Human hg19 genome assembly to the Chimpanzee panTro6 genome assembly from the UCSC genome browser (see Data availability). We converted the MAF formatted file (hg19.panTro6.synNet.maf.gz) to VCF format using MAFFilter v1.3.1^72^ (see Code availability) and extracted the chimpanzee allele at all variant sites in the UK Biobank.

### SNP annotations

We considered 165 single-SNP annotations (Supplementary Tables 2,3), including 163 annotations in the baseline-LF model^11^ and 2 annotations of CADD score^45^ for deleterious coding SNPs (common and low-frequency CADD score >20 SNPs, resp.). The 165 single-SNP annotations were constructed from 45 main functional annotations (baseline model version provided in Supplementary Table 2). Since we considered a different set of reference SNPs, we recomputed these main functional annotations. Specifically, the original .bed reference files were used for 36 main functional annotations. The annotations “Nucleotide diversity” and “Recombination rate” were recomputed following the original definition^8,11^. The annotation “MAF-adjusted LLD-AFR” was computed using the 1000 genome African population LD score^73^ (missing values imputed as 1). The annotations “Conserved (GERP RS ≥ 4)”, “Conserved (GERP NS)”, “CpG content”, “Deleterious (CADD ≥ 20)” were obtained from the CADD database v1.6^45^ (see Data availability). The annotations “Non-synonymous” and “Synonymous” were curated using SnpEff v4.3t^74^ (see Code availability). All single-SNP annotations analyzed are publicly available (see Data availability).

We constructed 136 SNP-pair annotations obtained by stratifying 34 main SNP-pair annotations by MAF (common or low-frequency) and LD (positive or negative): 3 proximity-based annotations (0-100bp, 100bp-1kb, 1-10kb), 5 gene-based annotations (e.g., same-gene promoter SNP pairs), 7 functional 0-100bp annotations, and 19 functional 0-1kb annotations (e.g., pairs of H3K27ac SNPs with distances 0-100bp) (Table 1, Supplementary Tables 4,5; Data availability). For gene-based annotations, we used GENCODE v41 for exon and gene annotations (Data availability) and downloaded the promoter annotation from ref.^70^, and annotated protein domains using VEP v102^75^ (Code availability). The functional SNP-pair annotations were constructed from 38 binary baseline model single-SNP functional annotations (Supplementary Table 2), restricted to functional SNP-pair annotations with at least 1 million SNP pairs (combined across MAF and LD bins). We excluded SNP-pair annotations involving one common SNP and one low-frequency SNP, because these SNP pairs had low levels of LD, limiting the informativeness of directional LD scores. All SNP-pair annotations analyzed are publicly available (see Data availability).

### Simulations

For all simulations, we used the UK Biobank genotype data of all 337,426 samples and all 1,161,341 SNPs on chromosome 1, analogous to previous work^8,11^. We considered two values of SCV (0.5 or 0.2) and two values of causal SNP proportion (0.2 or 0.1). We repeated all simulations 50 times. All simulation parameters are reported in Supplementary Table 7. We note that heritabilities are different from SCVs in causal simulations with nonzero SNP-pair effect correlations.

In null simulations, we simulated heritable traits with functional enrichment but zero SNP-pair effect correlations. First, we simulated per-SNP heritability of SNPs (Var(*β*_*i*_)) according to Equation (4), where we incorporated the LD-dependent genetic architecture by assigning nonzero *τ* to LD-related single-SNP annotations based on estimates from previous work^8,11^ and incorporated functional enrichments by assigning a positive *τ* to the common Super enhancer (Hnisz) single-SNP annotation, also motivated by previous work^5,8,11^ (Supplementary Table 7). Second, we simulated the MAF-dependent genetic architecture by further multiplying the simulated per-SNP heritability of each SNP *i* by [*p*_*i*_(1 − *p*_*i*_)]^(1+*α*)^, where *p*_*i*_ is the derived allele frequency and we used *α* = − 0.38 based on previous work^16^. Third, we simulated the sparse genetic architecture by randomly selecting a subset of causal SNPs, setting the simulated per-SNP heritability of non-causal SNPs to zero, and scaling up the simulated per-SNP heritability of causal SNPs to match the target SCV (making ∑_*i*_ Var(*β*_*i*_) equal to target SCV). Finally, we sampled causal SNP effect sizes for each SNP from a normal distribution with mean zero and variance equal to the simulated per-SNP heritability. We determined the true values of *τ* and *ω* by regressing simulated causal effects on the subset of causal single-SNP and SNP-pair annotations following Equation (4) and determined true values of other quantities based on true values of *τ* and *ω*.

In casual simulations, we simulated heritable traits with functional enrichment and nonzero SNP-pair effect correlations. In primary causal simulations, we simulated negative *ω* for positive-LD SNP pairs but zero *ω* for negative-LD SNP pairs, to mimic our findings in real-data analysis that positive-LD SNP pairs had strongly negative *ξ* estimates but negative-LD SNP pairs had very weakly positive *ξ* estimates (Figures 2, Supplementary Figure 8). First, we simulated LD-and-MAF dependent genetic architectures and functional enrichments for per-SNP heritability of SNPs by repeating the first and second steps in null simulations. Second, we assigned nonzero contributions to SNP-pair effect correlation (*ω*) to a subset of SNP-pair annotations (Supplementary Table 7) and calculated the correlation matrix of SNP effect sizes by summing up contributions from all causal SNP-pair annotations. Third, we calculated the covariance matrix of SNP effect sizes by scaling the simulated correlation matrix by simulated per-SNP heritability. Fourth, we simulated SNP causal effect sizes by blocks of 100 SNPs, randomly selecting a subset of blocks to be causal based on the target causal SNP proportion, and sampled causal SNP effect sizes from a multivariate normal distribution with zero mean and the simulated covariance matrix for causal SNP blocks (we removed negative eigenvalues from covariance matrices to keep them positive semidefinite). Fifth, we rescaled the simulated causal effect sizes to match the target SCV by scaling ∑_*i*_ Var(*β*_*i*_) to be equal to the target SCV. We calculated the true parameter values the same as in null simulations.

### Data analysis

We used genomic jackknife to assess standard error and statistical significance when aggregating dependent estimates, including analyses in Figure 4 and Supplementary Figure 9, and Supplementary Tables 15,17. For analysis of heterogeneity across diseases/traits (in *Analysis of 70 diseases and complex traits*), let *n* be the number of diseases/traits and let 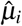 be the point estimate and SE of the *i*th trait. We assume that 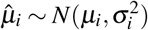. Let 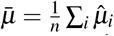be the unweighted mean. For the ratio between across-trait variance and average SE, the across-trait variance is estimated as ^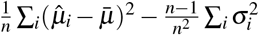^ (second term corrects for bias), and the average SE is computed as ^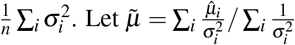^. be the weighted mean. The chi-square statistic is 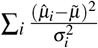 and follows a 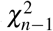 distribution.

### Forward evolutionary simulations

Forward evolutionary simulations were performed on SLiM v3.6^76^ (Code availability) using a fixed population size of 10,000 diploid individuals, each with a single chromosome of length 100kb, mutation rate *μ* = 1 *×*10^−8^, and recombination rate 1 *×*10^−8^. For simulations of stabilizing selection, new mutations were introduced at rate *μ* with effect sizes of −*β* (trait-decreasing), 0 (neutral), or +*β* (trait-increasing) (with equal probability); *β* = 0.1 was used in the main simulation, and additional *β* values were considered for simulation of linkage disequilibrium varying over a log-scaled range from 1 *×*10^−4^ to 1 (Supplementary Figure 12b). At the end of each generation, aggregate trait effect *g* for each individual was calculated as 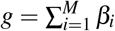 across *M* variants each with effect size *β*_*i*_. Individual fitness *W* (*g*) (as a function of aggregate trait effect *g* for each individual in a given generation) was calculated as 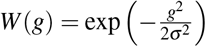 depending on the width of fitness function parameter *σ*, following ref.^38^. We considered 3 values for the width parameter: strong selection (*σ* = 2, used in the main simulation), moderate selection (*σ* = 4), and effectively neutral (*σ* = 1*×* 10^6^) (Supplementary Figure 12a). Simulations were run for 10*N* = 100, 000 generations. Pairwise linkage disequilibrium *D* was computed using emeraLD^77^ v0.1 (Supplementary Figure 12) (Code availability), or using correlation coefficient (Figure 6). An aggregate of 5,000 simulated populations was run, and the mean statistic (e.g., *ξ* or *D*) was summarized within each run and then between runs to derive mean values and confidence intervals.

## Data availability

Information of imputed SNPs and corresponding ancestral alleles, GWAS summary statistics, baseline-SP single-SNP and SNP-pair annotations, LD scores, directional LD scores, and LDSPEC output from this study are available at https://figshare.com/projects/LD_SNP-pair_effect_correlation_regression_LDSPEC_/188052. We did not release in-sample LD files due to their large sizes; similar in-sample LD files can be found in ref.^43^. The whole genome alignment of the Human hg19 genome assembly to the Chimpanzee panTro6 genome assembly is available at http://hgdownload.cse.ucsc.edu/goldenpath/hg19/vsPanTro6/. CADD database v1.6^45^ is available at https://cadd.gs.washington.edu/download. GENCODE v41 is available at https://www.gencodegenes.org/human/release_41.html. The promoter annotation from ref.^70^ is available at https://alkesgroup.broadinstitute.org/cS2G.

## Code availability

Software implementing LDSPEC and code for generating all results of the paper are available at https://github.com/martinjzhang/LDSPEC. MAFFilter v1.3.1^72^ is available at https://jydu.github.io/maffilter/. SnpEff v4.3t^74^ is available at http://pcingola.github.io/SnpEff/. VEP v102^75^ is available at https://useast.ensembl.org/info/docs/tools/vep/script/vep_download.html. SLiM version v3.6 is available at https://github.com/MesserLab/SLiM/releases/tag/v3.6. emeraLD version v0.1 is available at https://github.com/statgen/emeraLD.

## Supplementary Note

### 1 Proofs and derivations

#### 1.1 Derivation of the regression equation

Regression equation:

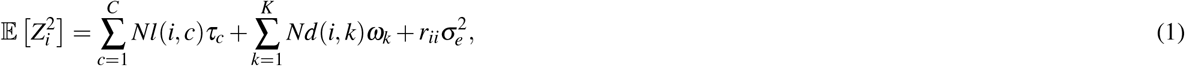

where 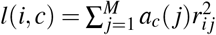 and 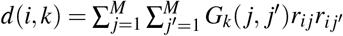.

*Proof*. Let **X**_*i*_ = [*X*_1*i*_, *· · ·, X*_*Ni*_]^*T*^ be the *i*-th column of **X**. The summary association statistic for SNP *i* satisfies

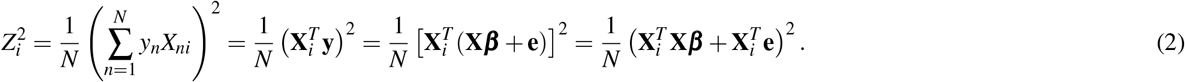

Next, the expectation of 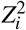 can be written as

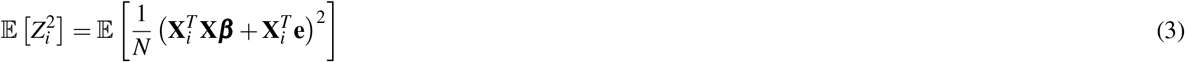

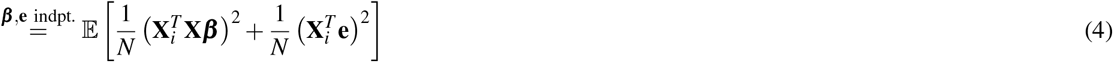

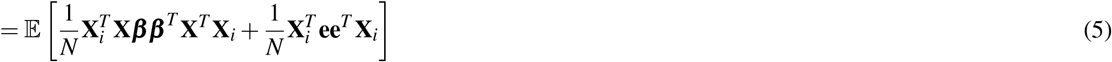

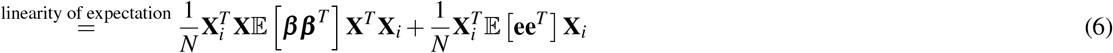

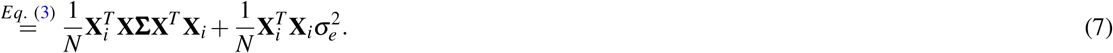

Let 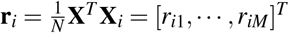 be the signed LD between SNP *i* and other SNPs. We have

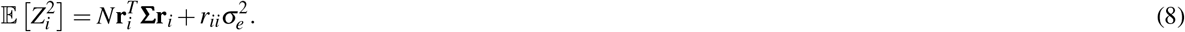

Define the vector form of single-SNP annotation **a**_*c*_ = [*a*_*c*_(1),*· · ·, a*_*c*_(*M*)]^*T*^ and the matrix form of SNP-pair annotation **G**_*k*_ : [**G**_*k*_]_*i j*_ = *G*_*k*_(*i, j*). Taking Eq. (4) into the above equation to have

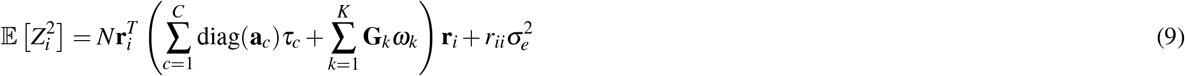

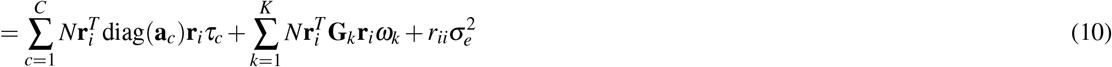

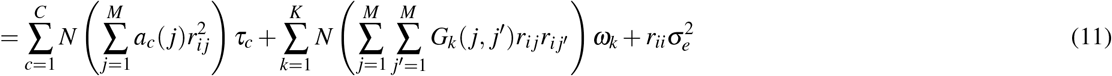

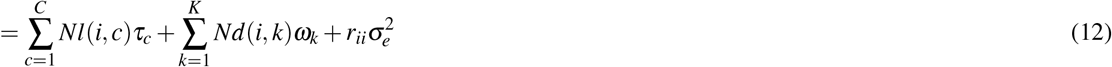

#### 1.2 Derivation of heritability

Heritability for binary single-SNP annotation *c*:

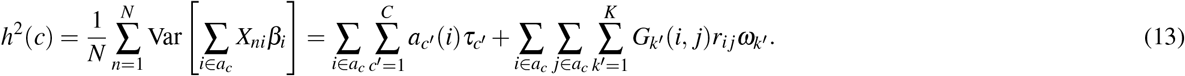

*Proof*. Let **X**_*n*_ = [*X*_*n*1_,, *X*_*nM*_]^*T*^ be the *n*-th row of **X**. Let 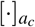 denote restricting the corresponding vector/matrix to elements in *a*_*c*_.

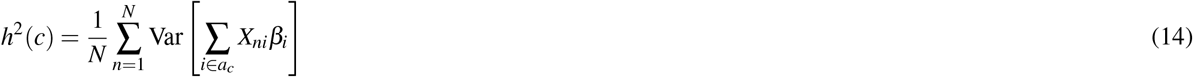

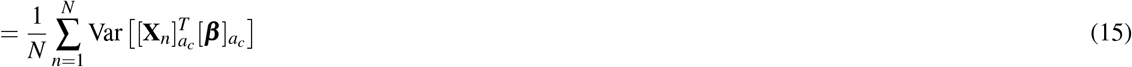

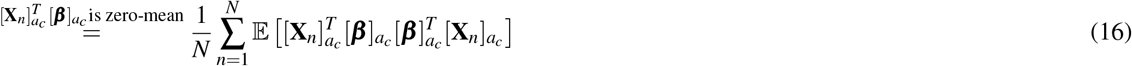

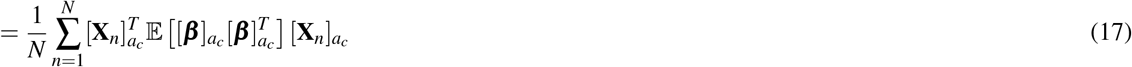

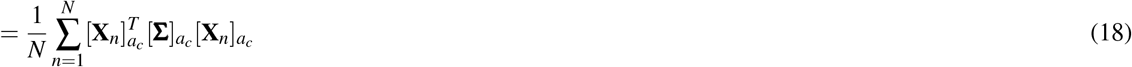

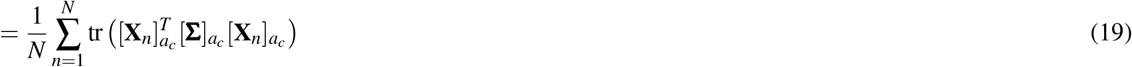

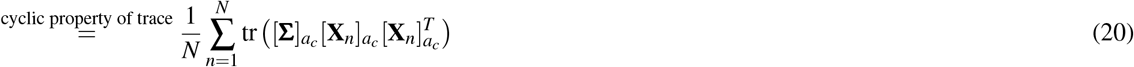

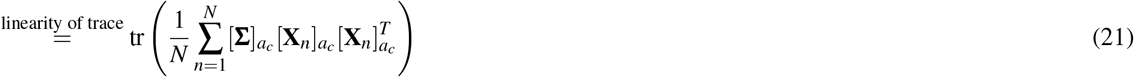

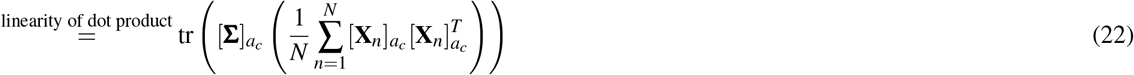

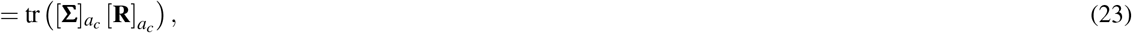

where for the last equation, we note that ^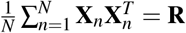^, the LD matrix whose *i j*-th element is equal to *r*_*i j*_.

Define the vector form of single-SNP annotation **a**_*c*_ = [*a*_*c*_(1), *· · ·, a*_*c*_(*M*)]^*T*^ and the matrix form of SNP-pair annotation **G**_*k*_ : [**G**_*k*_]_*i j*_ = *G*_*k*_(*i, j*). Furthermore,

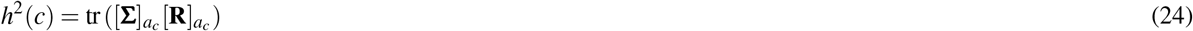

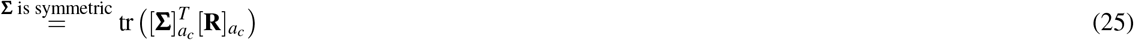

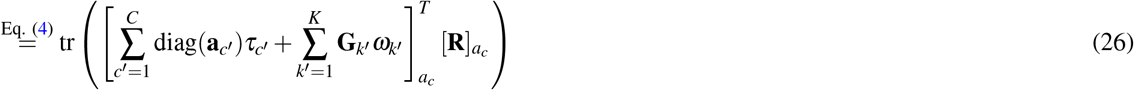

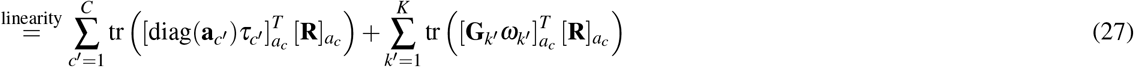

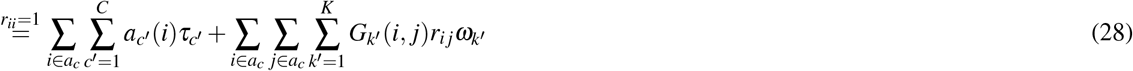

## Supplementary Tables

See Supplementary Excel file

**Supplementary Table 1. GWAS diseases and complex traits**. We report the name, identifier, indication of 29 independent traits, and number of samples for 70 diseases/traits analyzed in the paper. For each disease/trait, we also report estimates of heritability, heritability SE, and z-score for nonzero heritability from LDSPEC with the baseline-SP model.

See Supplementary Excel file

**Supplementary Table 2. Main single-SNP annotations**. We report the name, identifier, type, number of common SNPs (MAF ≥5%), number of low-frequency SNPs (0.5% ≤MAF*<*5%), reference, source, and version of baseline model for main 45 single-SNP annotations in the baseline-SP model.

See Supplementary Excel file

**Supplementary Table 3. Single-SNP annotations in baseline-SP**. We report the name, type, and number of SNPs for the 165 single-SNP annotations in the baseline-SP model.

See Supplementary Excel file

**Supplementary Table 4. Main SNP-pair annotations**. We report the name, identifier, type, and description for 34 main SNP-pair annotations in the baseline-SP model. For each SNP-pair annotation, we also report number of SNP pairs and average distance (combined across common negative-LD, low-frequency negative-LD, common positive-LD, low-frequency positive-LD), and we report average LD (for common negative-LD, low-frequency negative-LD, common positive-LD, low-frequency positive-LD, separately).

See Supplementary Excel file

**Supplementary Table 5. SNP-pair annotations in baseline-SP**. We report the name and number of SNP pairs for the 136 SNP-pair annotations in the baseline-SP model.

See Supplementary Excel file

**Supplementary Table 6. Correlation of LD and directional LD scores**. We report the correlation across 14,820,648 SNPs between 165 single-SNP annotations and 136 SNP-pair annotations in the baseline-SP model.

See Supplementary Excel file

**Supplementary Table 7. Simulation parameters**. We report the simulation name, term, and values for all simulations performed in the paper. “h2g” denotes target SCV (may be different from heritability in causal simulations), “p_causal” denotes proportion of causal SNPs, and “alpha” denotes the MAF-dependent genetic architecture, i.e., scaling the per-SNP heritability by [MAF(1 − MAF)] ^(1+*α*)^.

See Supplementary Excel file

**Supplementary Table 8. Numerical results for null simulations in Figure 1a**. We report the annotation name, term, term identifier, and true value for all estimates across the 165 single-SNP annotations and 136 SNP-pair annotations in the baseline-SP model. For each term and each annotation, we report LDSPEC estimates aggregated across the 50 simulation replicates: jackknife 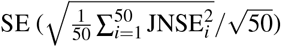, empirical mean (mean across 50 estimates), empirical SE (SD across 50 estimates divided by 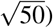), empirical p-value (assuming normal distribution), and empirical FWER (*P*<0.05/165 for single-SNP annotations and *P*<0.05/136 for SNP-pair annotations).

See Supplementary Excel file

**Supplementary Table 9. Numerical results for causal simulations in Figure 1b**. We report the annotation name, term, term identifier, and true value for all estimates across the 165 single-SNP annotations and 136 SNP-pair annotations in the baseline-SP model. For each term and each annotation, we report LDSPEC estimates aggregated across the 50 simulation replicates: jackknife 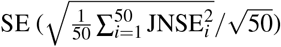, empirical mean (mean across 50 estimates), empirical SE (SD across 50 estimates divided by 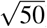), empirical p-value (assuming normal distribution), and empirical FWER (*P*<0.05/165 for single-SNP annotations and *P*<0.05/136 for SNP-pair annotations).

See Supplementary Excel file

**Supplementary Table 10. LDSPEC results for single-SNP annotations and 70 diseases/traits**. We report trait identifier, annotation identifier, annotation type, and number of SNPs for 165 single-SNP annotations. We report point estimates, SE, and p-values of *τ*, heritability, SCV, heritability enrichment, and heritability shrinkage for 165 single-SNP annotations and 70 diseases/traits.

See Supplementary Excel file

**Supplementary Table 11. LDSPEC results for SNP-pair annotations and 70 diseases/traits**. We report trait identifier, annotation identifier, and number of SNP pairs for 136 SNP-pair annotations. We report point estimates, SE, and p-values of *ω*, total SNP-pair effect covariance, *ξ*, total excess SNP-pair effect covariance, and *ξ*^∗^ for 70 diseases/traits and 136 SNP-pair annotations.

See Supplementary Excel file

**Supplementary Table 12. Meta-analyzed LDSPEC results for single-SNP annotations**. We report meta-analyzed point estimates, SE, and p-values of *τ*, heritability, SCV, heritability enrichment, and heritability shrinkage for 165 single-SNP annotations. The meta-analysis was performed across 29 independent diseases/traits.

See Supplementary Excel file

**Supplementary Table 13. Meta-analyzed LDSPEC results for SNP-pair annotations**. We report meta-analyzed point estimates, SE, and p-values of *ω*, total SNP-pair effect covariance, *ξ*, total excess SNP-pair effect covariance, and *ξ*^∗^ for 136 SNP-pair annotations. The meta-analysis was performed across 29 independent diseases/traits.

See Supplementary Excel file

**Supplementary Table 14. Numerical results for Figure 2**. We report annotation name, *ξ* estimate, SE of *ξ* estimate, p-value of *ξ* estimate, and FWER of *ξ* estimate for low-frequency negative-LD, common negative-LD, low-frequency positive-LD, and common positive-LD SNP-pair annotations in Figure 2, respectively.

See Supplementary Excel file

**Supplementary Table 15. Jacknife-estimated differences for comparisons in Figure 2**. We report annotation name, first stratum, second stratum, estimated difference, SE, p-value, and FWER for each comparison.

See Supplementary Excel file

**Supplementary Table 16. Numerical results for Figure 3**. We report annotation name, *ξ*^∗^ estimate, SE of *ξ*^∗^ estimate, p-value of *ξ*^∗^ estimate, and FWER of *ξ* estimate for the 0-100bp and 0-1kb common positive-LD, low-frequency positive-LD, common negative-LD, and low-frequency negative-LD SNP-pair annotations, respectively.

See Supplementary Excel file

**Supplementary Table 17. Jacknife-estimated differences for comparisons in Figure 3**. We report annotation name, first stratum, second stratum, estimated difference, SE, p-value, and FWER for each comparison.

See Supplementary Excel file

**Supplementary Table 18. Numerical results for Figure 4**. We report the annotation name, heritability enrichment estimate, SE of heritability enrichment estimate, *ξ*^∗^ estimate, and SE of *ξ*^∗^ estimate for SNP-pair annotations in Figure 4.

See Supplementary Excel file

**Supplementary Table 19. Heterogeneity across traits**. We report the chi-square statistic, p-value, within-trait variance, between-trait variance, variance ratio (within over between), FWER (across 136 SNP-pair annotations tested), and FDR (across 136 SNP-pair annotations tested).

See Supplementary Excel file

**Supplementary Table 20. Numerical results for Figure 5**. We report the trait name, heritability estimate, SE of heritability estimate, SCV estimate, SE of SCV estimate, heritability shrinkage estimate, and SE of heritability shrinkage estimate for 70 diseases/traits.

See Supplementary Excel file

**Supplementary Table 21. Numerical results for Figure 6**. We report the distance bin, MAF bin, LD bin, *ξ* estimate, and SE of *ξ* estimate for SNP-pair categories in Figure 6.

## Supplementary Figures

**Supplementary Figure 1.**
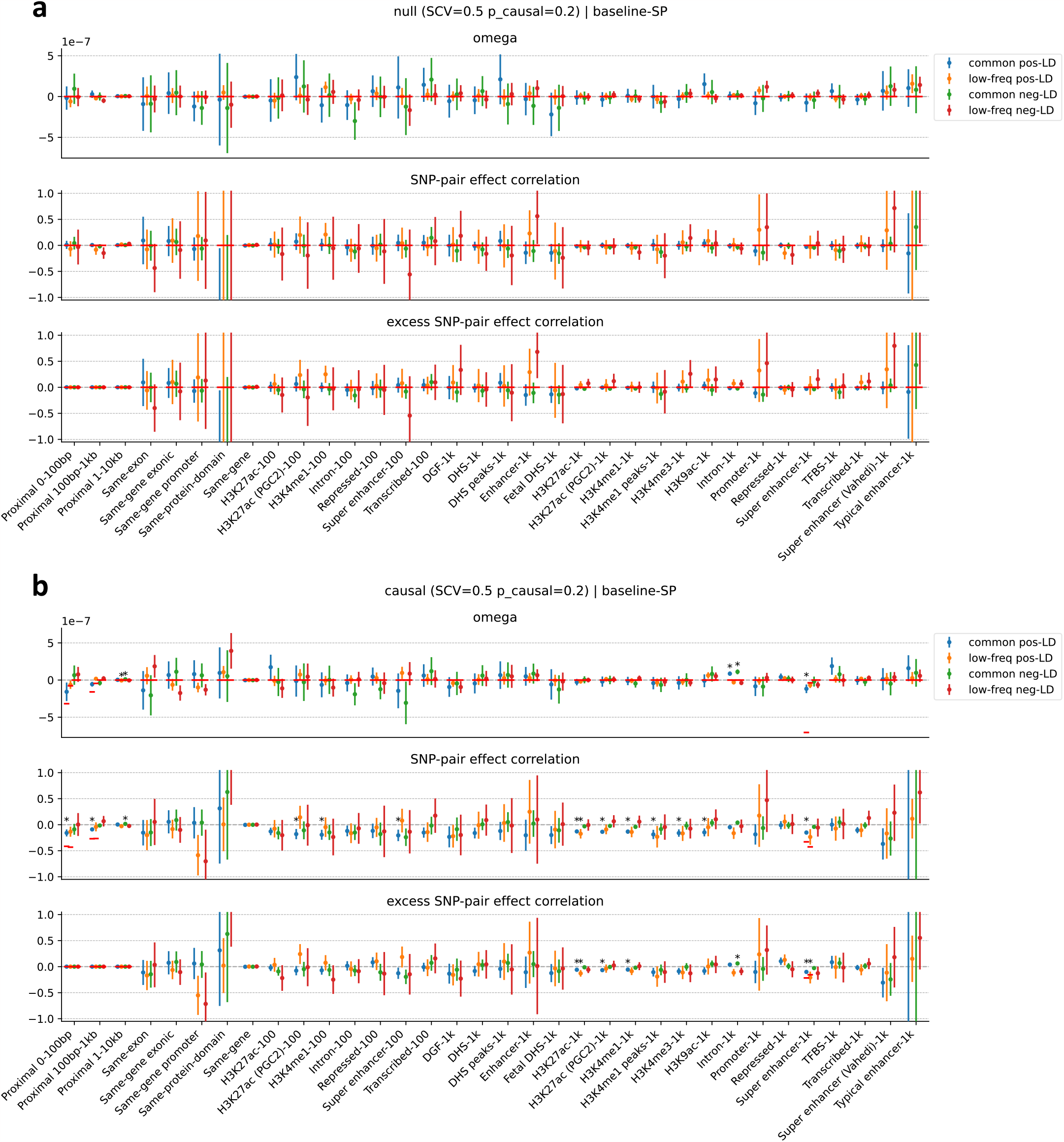
Additional results of estimates for SNP-pair annotations in null and casual simulations in Figure 1. We report estimates of *ω, ξ*, and *ξ*^∗^ for 136 SNP-pair annotations in the baseline-SP model. **(a)** for null simulations and **(b)** for causal simulations. Error bars denote 95% confidence intervals around the mean of 50 simulation replicates; “*” denotes statistical significance after multiple testing correction (*P*<0.05/136). Red horizontal lines represent the true simulated values for SNP-pair annotations whose true values are available.

**Supplementary Figure 2.**
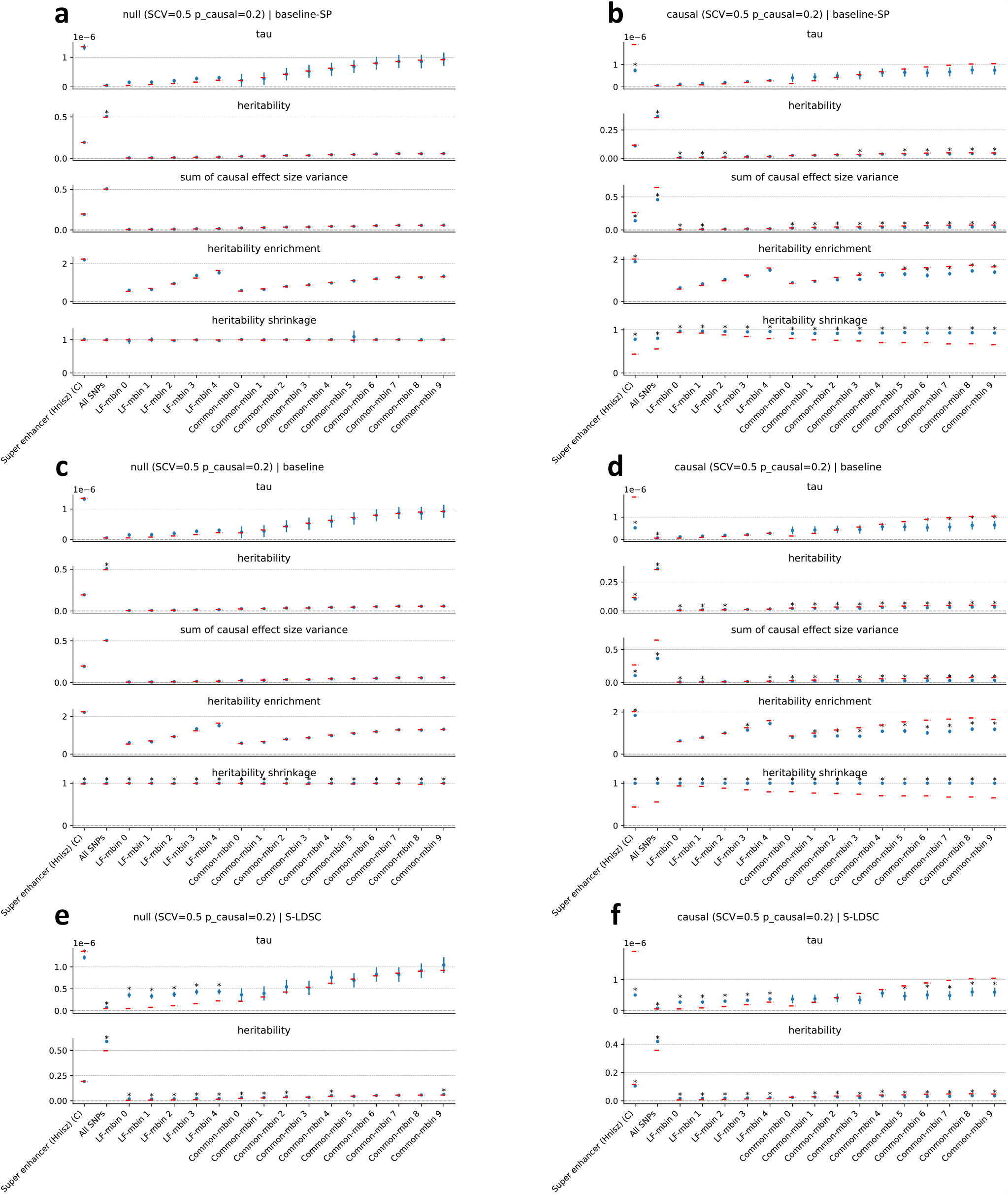
Additional results of estimates for single-SNP annotations in null and casual simulations in Figure 1. We report estimates of *τ*, heritability, SCV, heritability enrichment, and heritability shrinkage for 17 binary single-SNP annotations whose true values for all 5 terms are available. **(a)** for null simulations and **(b)** for causal simulations. We also report the corresponding estimates using LDSPEC + baseline (without SNP-pair annotations) in panels c,d, and corresponding estimates (*τ* and heritability) using S-LDSC^1^ + baseline (without SNP-pair annotations) in panels e,f. Error bars denote 95% confidence intervals around the mean of 50 simulation replicates; “*” denotes statistically significantly different from the true values after multiple testing correction (*P*<0.05/165). Red horizontal lines represent the true values.

**Supplementary Figure 3.**
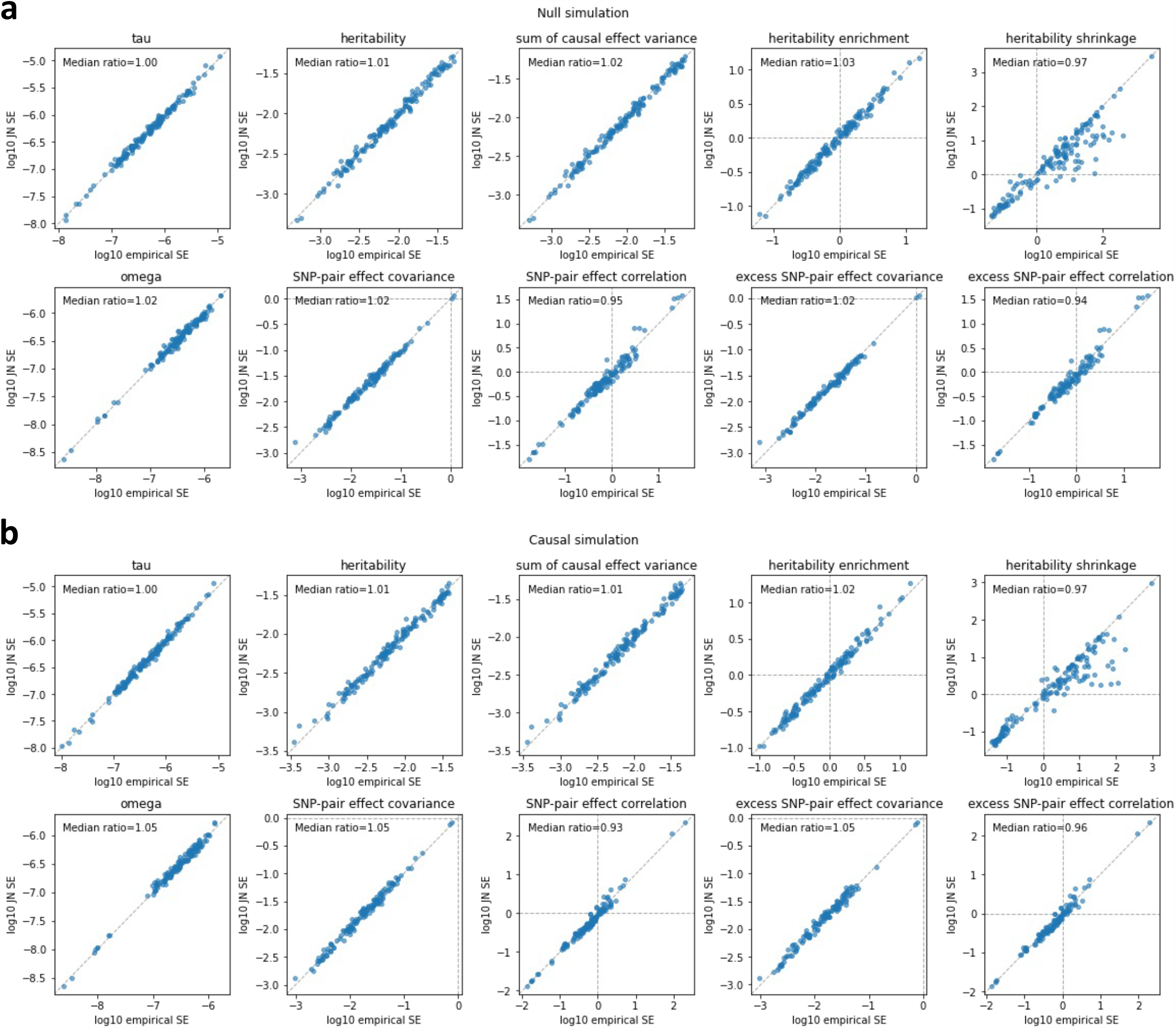
Calibration of CIs for null and causal simulations in Figure 1. Results are shown for estimates of *τ*, heritability, SCV, heritability enrichment, heritability shrinkage, *ω*, SNP-pair effect covariance, *ξ*, excess SNP-pair effect covariance, and *ξ*^∗^, respectively. **(a)** for null simulations and **(b)** for causal simulations. Each point represents an annotation, x-axis represents the log 10 empirical SE (SD of estimates across simulation replicates), and y-axis represents the log 10 jackknife 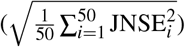. The median of ratios between jackknife SE and empirical SE across annotations is provided in the figure annotation. We note that the p-value of *ξ* is based on estimates of SNP-pair effect covariance, and the p-value of *ξ*^∗^ is based on estimates of excess SNP-pair effect covariance (Methods).

**Supplementary Figure 4.**
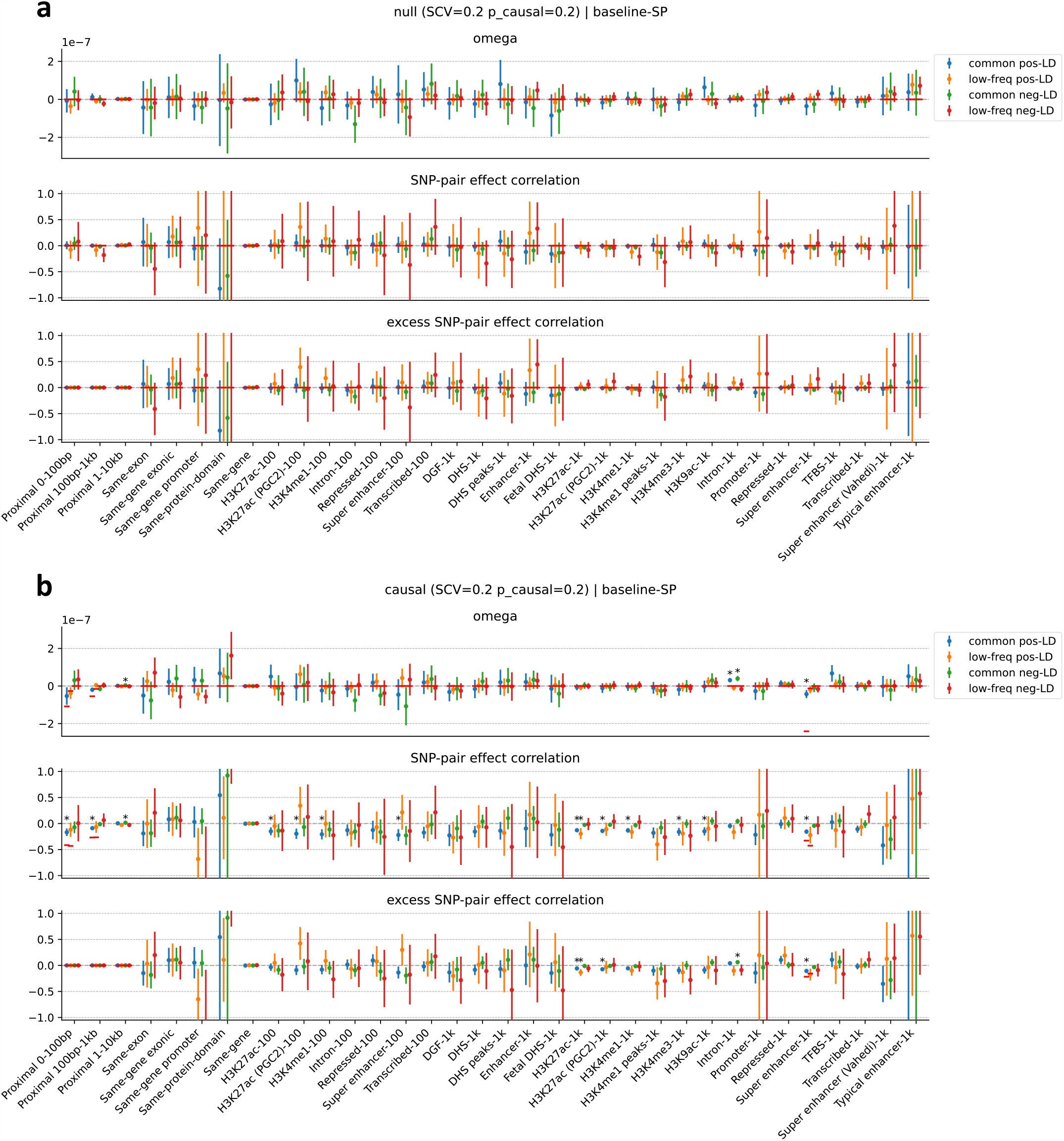
Null and causal simulations with a lower value of SCV. **(a)** Null simulations with SCV of 0.2 (instead of 0.5) and causal SNP proportion of 0.2. **(b)** Causal simulations with SCV of 0.2 (instead of 0.5) and causal SNP proportion of 0.2. We report estimates of *ω, ξ*, and *ξ*^∗^ for 136 SNP-pair annotations in the baseline-SP model. Error bars denote 95% confidence intervals around the mean of 50 simulation replicates; “*” denotes statistical significance after multiple testing correction (*P*<0.05/136). Red horizontal lines represent the true simulated values for SNP-pair annotations whose true values are available.

**Supplementary Figure 5.**
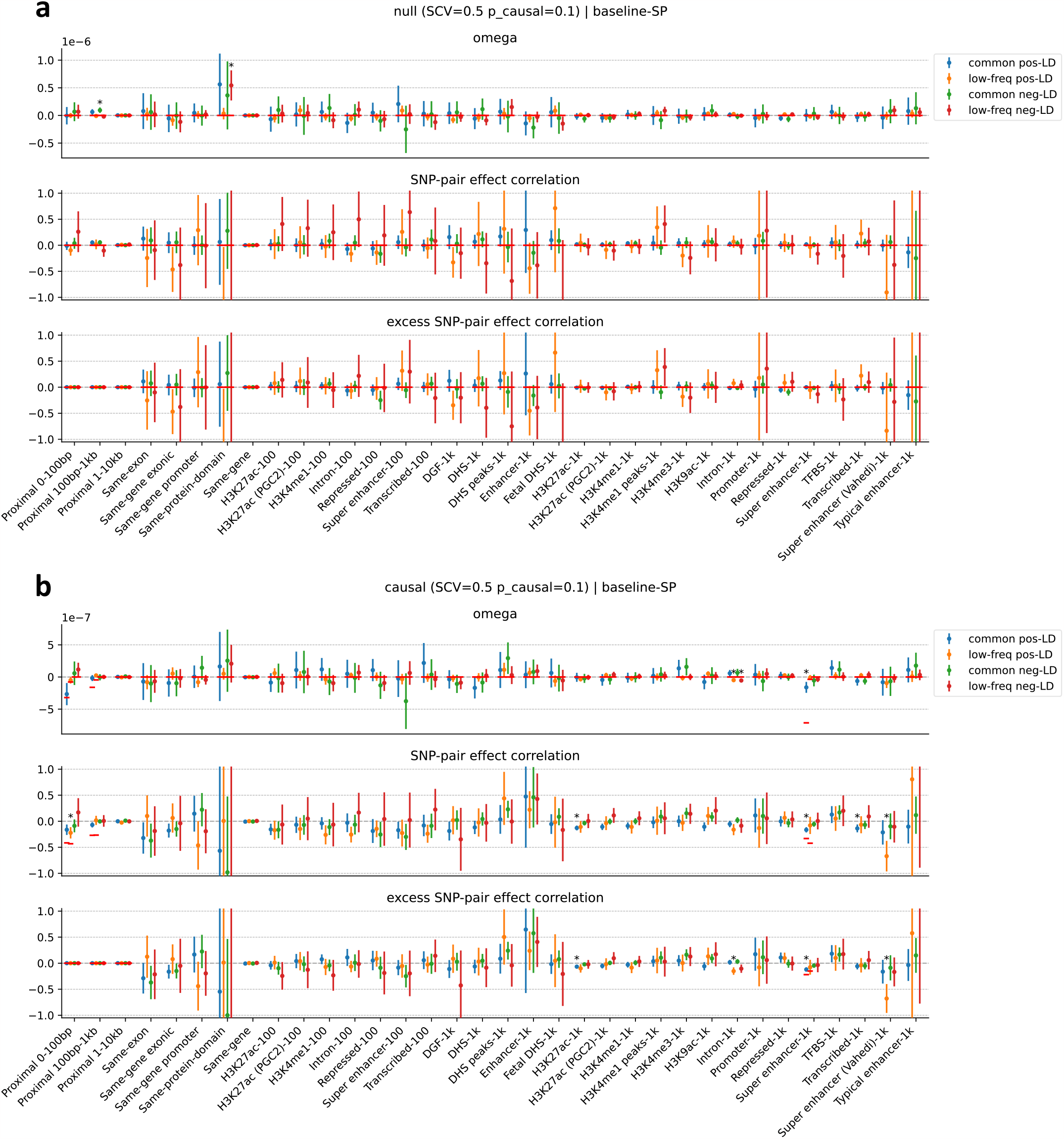
Null and causal simulations with a lower value of causal SNP proportion. **(a)** Null simulations with SCV of 0.5 and causal SNP proportion of 0.1 (instead of 0.2). **(b)** Causal simulations with SCV of 0.5 and causal SNP proportion of 0.1 (instead of 0.2). We report estimates of *ω, ξ*, and *ξ*^∗^ for 136 SNP-pair annotations in the baseline-SP model. Error bars denote 95% confidence intervals around the mean of 50 simulation replicates; “*” denotes statistical significance after multiple testing correction (*P*<0.05/136). Red horizontal lines represent the true simulated values for SNP-pair annotations whose true values are available.

**Supplementary Figure 6.**
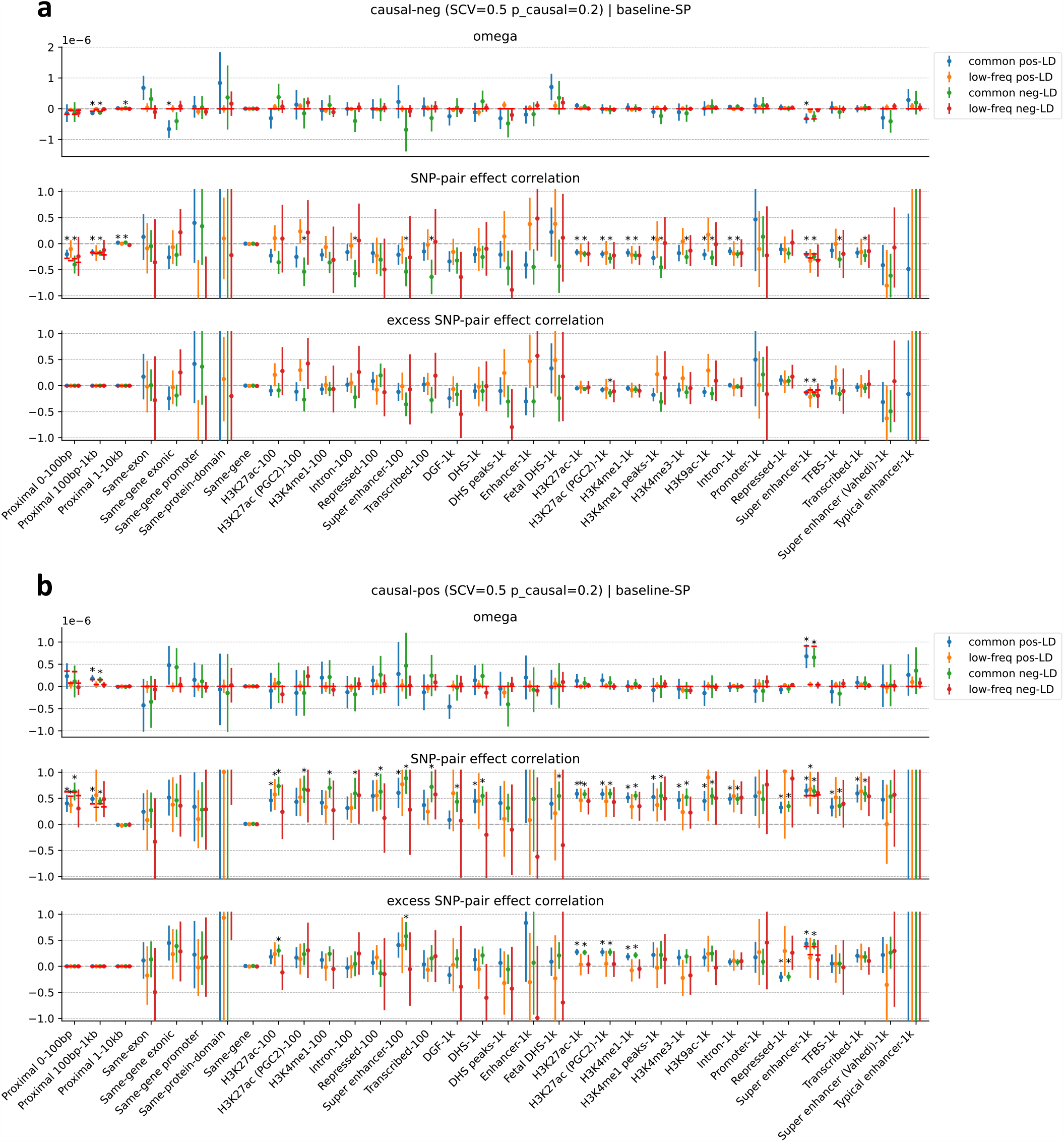
Non-LD-stratified causal simulations. **(a)** Causal simulations with negative *ω* for both positive-LD and negative-LD SNP-pair annotations (vs. negative *ω* for only positive-LD SNP-pair annotations in primary simulations), SCV of 0.5, and causal SNP proportion of 0.2. **(b)** Causal simulations with positive *ω* for both positive-LD and negative-LD SNP-pair annotations (vs. negative *ω* for only positive-LD SNP-pair annotations in primary simulations), SCV of 0.5, and causal SNP proportion of 0.2. We report estimates of *ω, ξ*, and *ξ*^∗^ for 136 SNP-pair annotations in the baseline-SP model. Error bars denote 95% confidence intervals around the mean of 50 simulation replicates; “*” denotes statistical significance after multiple testing correction (*P*<0.05/136). Red horizontal lines represent the true values for SNP-pair annotations whose true values are available.

**Supplementary Figure 7.**
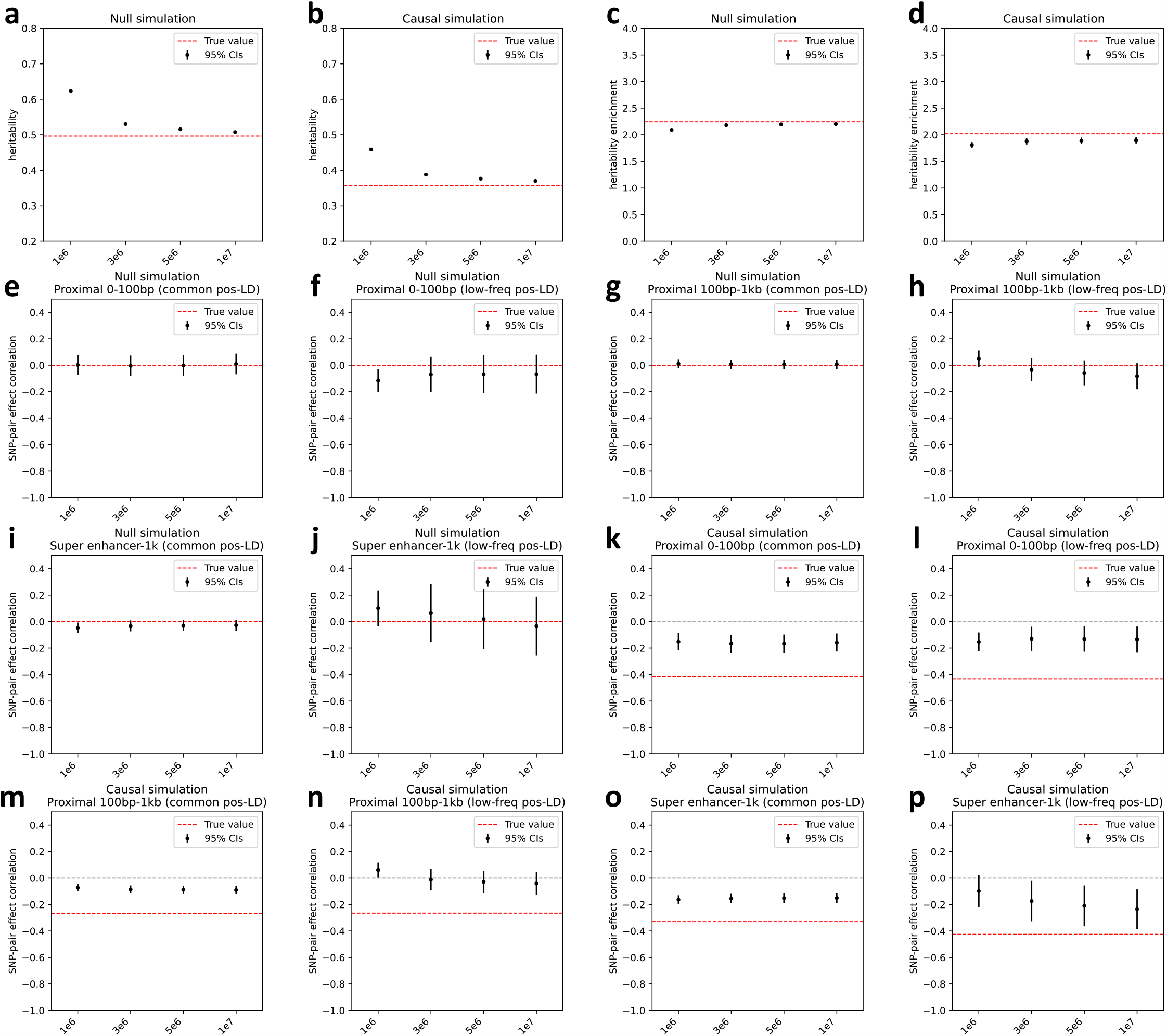
Results for applying LDSPEC to the primary null and causal simulation data with LD and directional LD scores computed with smaller window sizes. We considered 3 smaller window sizes: 1Mb, 3Mb, 5Mb (instead of 10Mb). **(a-b)** Estimates of heritability in null and causal simulations. **(c-d)** Estimates of heritability enrichment for the common Super enhancer (Hnisz) annotation in null and causal simulations (simulated to have a positive *τ* in both null and causal simulations). **(e-p)** Estimates of *ξ* in null and causal simulations for the 6 SNP-pair annotations simulated to have negative *ω* in the causal simulation. Error bars denote 95% confidence intervals around the mean of 50 simulation replicates. Red horizontal lines represent the true simulated values.

**Supplementary Figure 8.**
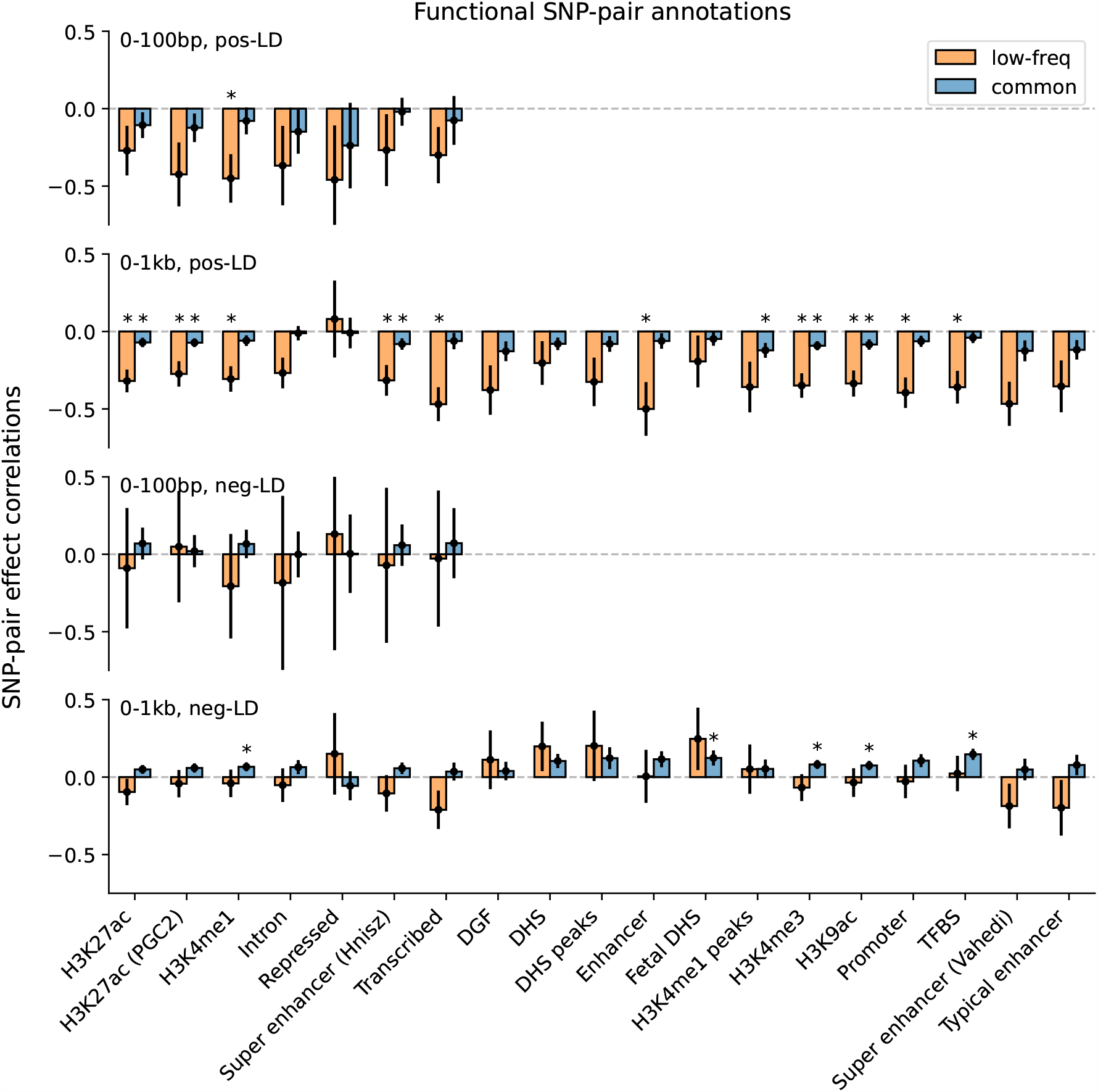
Estimates of SNP-pair effect correlation (*ξ*) across 29 independent diseases and complex traits for functional SNP-pair annotations. We report meta-analyzed *ξ* estimates across 29 independent diseases for 7 functional 0-100bp and 19 functional 0-1kb SNP-pair annotations. Results are shown for the positive-LD 0-100bp, positive-LD 0-1kb, negative-LD 0-100bp, and negative-LD 0-1kb SNP-pair annotations in the 4 panels, respectively, and are stratified by MAF in each panel. Error bars denote 95% confidence intervals. “*” denotes statistical significance after multiple testing correction across estimates on the figure (*P*<0.05/136).

**Supplementary Figure 9.**
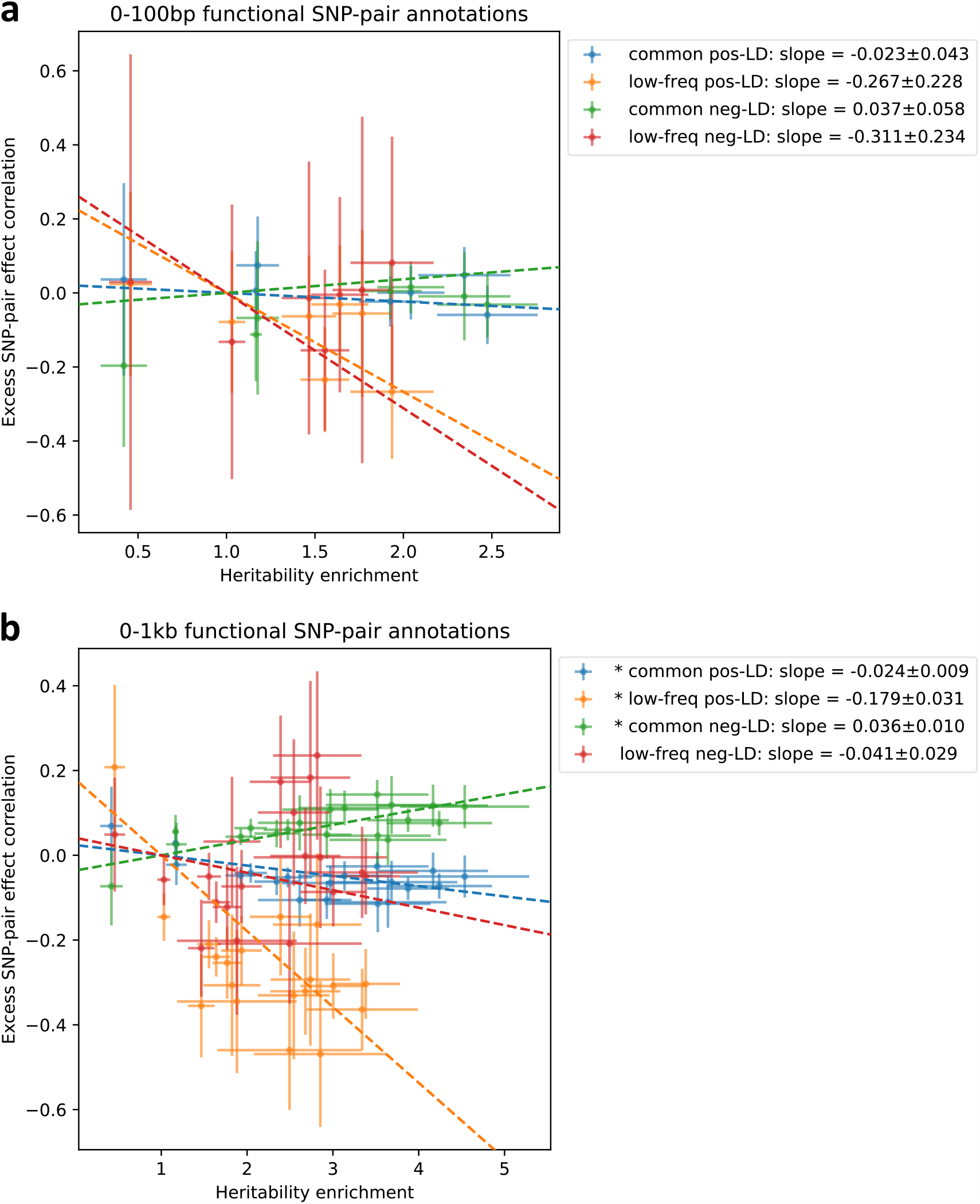
Comparison between estimates of heritability enrichment and estimates of excess SNP-pair effect correlation estimate (*ξ*^∗^) across functional SNP-pair annotations. Panels a and b show results for functional 0-100bp and functional 0-1kb SNP-pair annotations, respectively. Each dot represents a SNP-pair annotation, x-axis represents the meta-analyzed estimate of heritability enrichment, and y-axis represents the meta-analyzed estimate of *ξ*^∗^ (across 29 independent diseases/traits). In each panel, results are shown for the common positive-LD, low-frequency positive-LD, common negative-LD, and low-frequency positive-LD SNP-pair annotations separately. Error bars denote 95% confidence intervals. Regression slopes are provided with SEs in the figure legend; “*” denotes statistical significance after multiple testing correction (*P*<0.05/4).

**Supplementary Figure 10.**
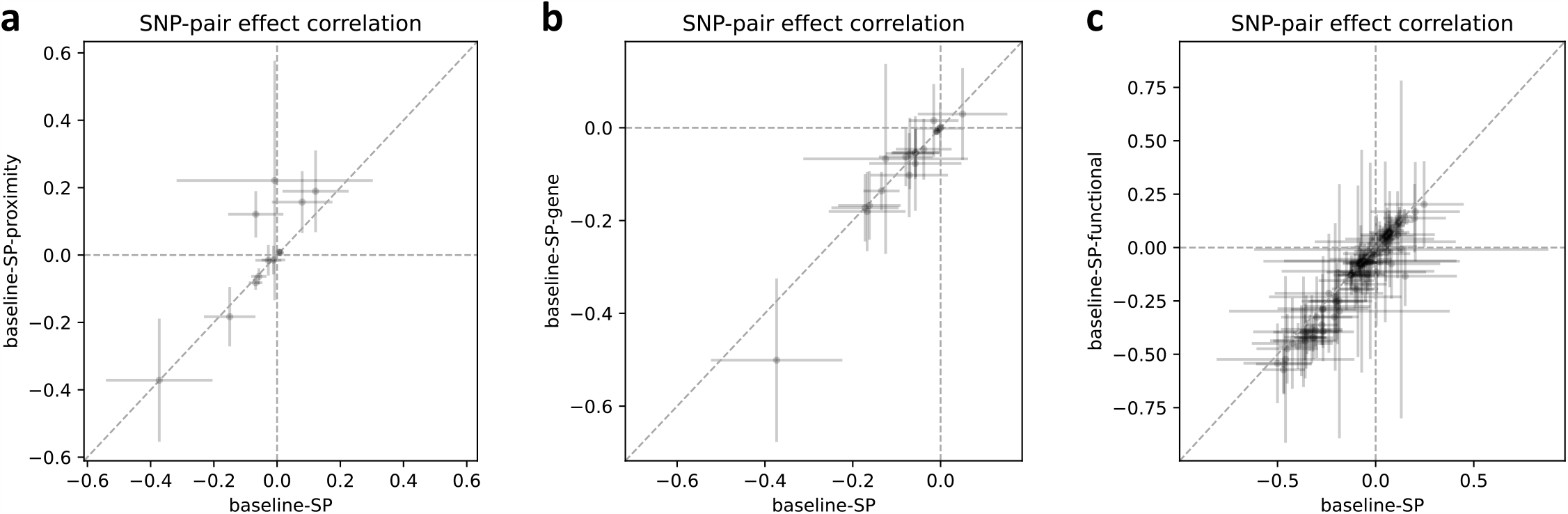
Comparison of *ξ* estimates of LDSPEC with alternative heritability models. **(a)** Comparison of meta-analyzed *ξ* estimates between the baseline-SP model (x-axis) and the baseline-SP-proximity model for 12 proximity-based SNP-pair annotations shared between the two models. **(b)** Comparison of meta-analyzed *ξ* estimates between the baseline-SP model (x-axis) and the baseline-SP-gene model for 20 gene-based SNP-pair annotations shared between the two models. **(c)** Comparison of meta-analyzed *ξ* estimates between the baseline-SP model (x-axis) and the baseline-SP-functional model for 104 functional SNP-pair annotations shared between the two models. Each dot represents a SNP-pair annotation. No difference between the x-value and y-value is statistically significant (*P*>0.05/136).

**Supplementary Figure 11.**
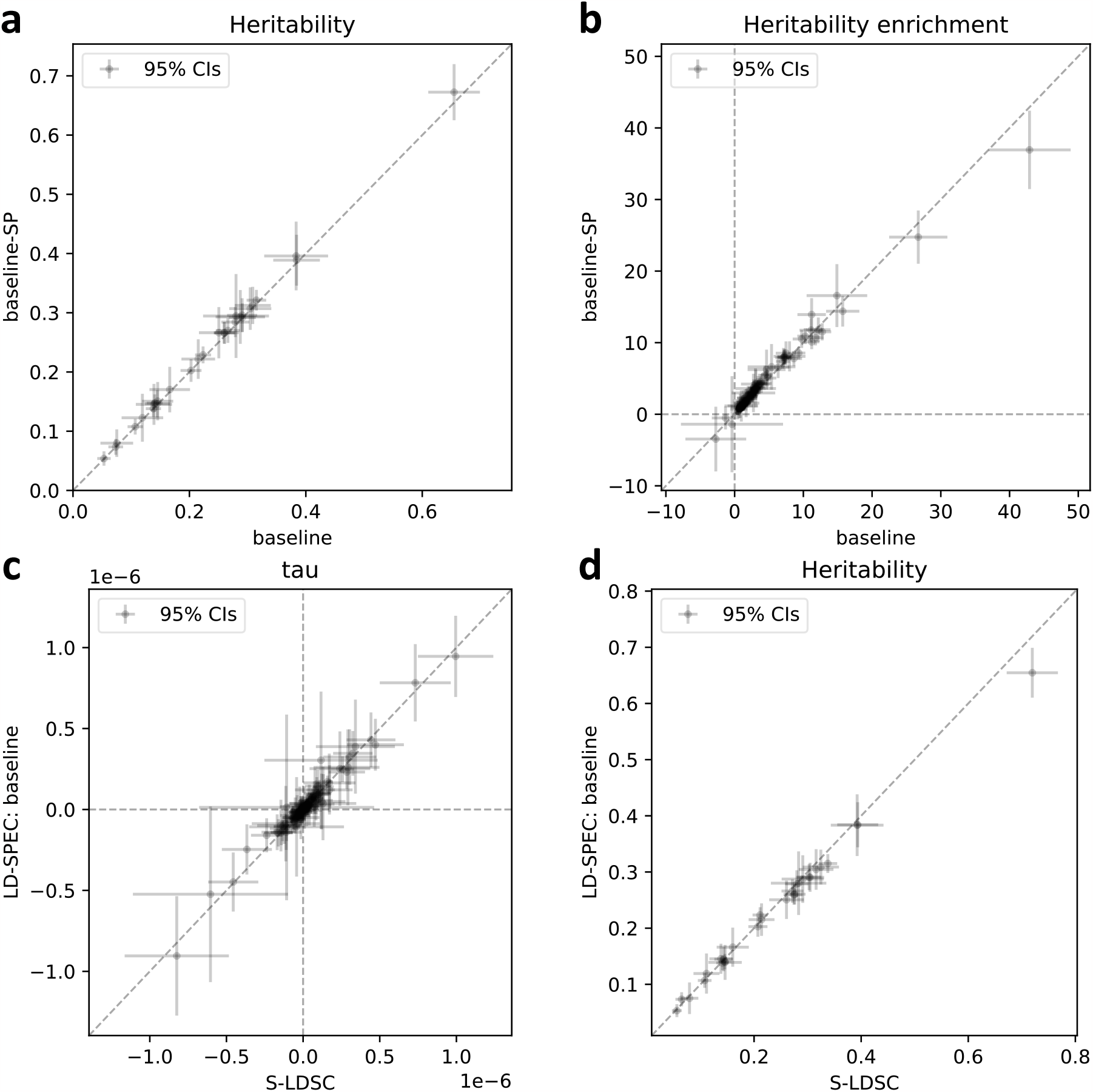
Comparison of estimates for single-SNP annotations. **(a)** Comparison of heritability estimates using LDSPEC with the baseline model (x-axis) and the baseline-SP model (y-axis) for 29 independent diseases/traits. **(b)** Comparison of meta-analyzed heritability enrichment estimates using LDSPEC with the baseline model (x-axis) and the baseline-SP model (y-axis) for 165 single-SNP annotations. **(c)** Comparison of meta-analyzed *τ* estimates between S-LDSC^1^ (x-axis) and LDSPEC (y-axis) (both using the baseline model) for 165 single-SNP annotations. **(d)** Comparison of heritability estimates between S-LDSC^1^ (x-axis) and LDSPEC (y-axis) (both using the baseline model) for 29 independent diseases/traits. No difference between the x-value and y-value is statistically significant (*P*>0.05/29 for panels a,d, *P*>0.05/165 for panels b,c).

**Supplementary Figure 12.**
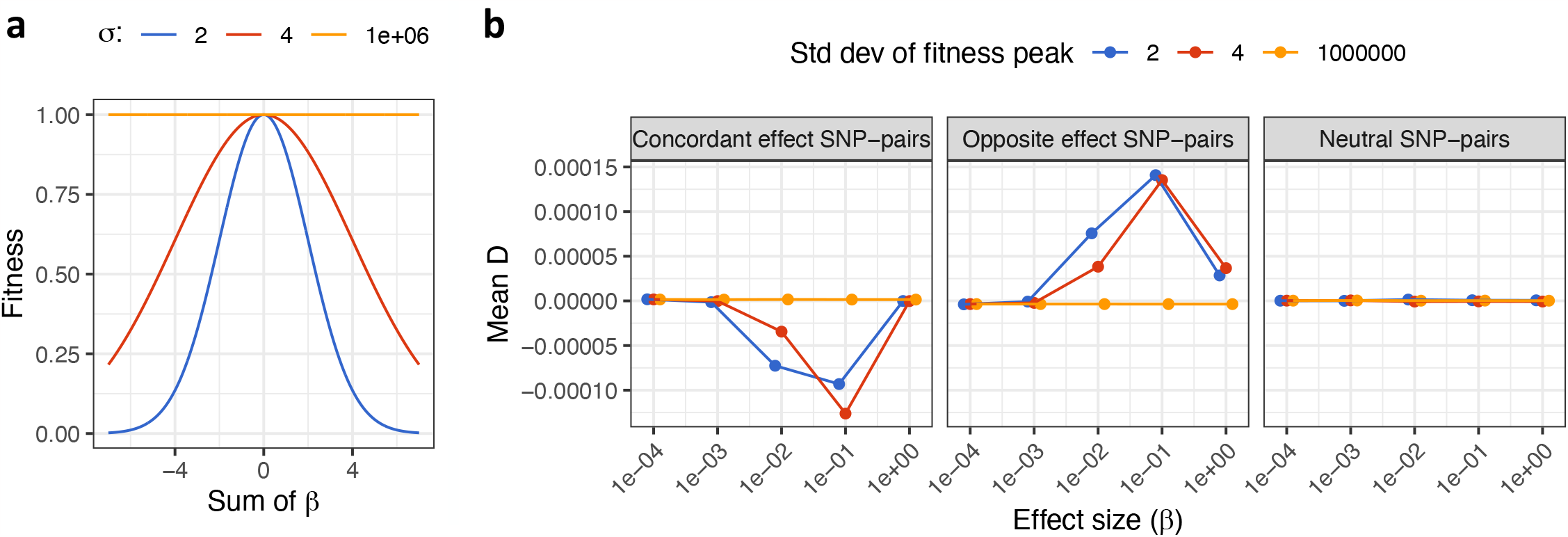
Additional results for forward evolutionary simulations. **(a)** Fitness as a function of sum of causal effects (aggregate SNP effect on trait) under a stabilizing selection model. The 3 curves correspond to strong selection (width *σ* = 2), moderate selection (width *σ* = 4), and no selection (width *σ* = 1 *×* 10^6^). **(b)** LD measured by *D* as a function of effect size (*β*) for concordant-effect SNP pairs (left), opposite-effect SNP pairs (middle), and neutral SNP pairs (right, at least one zero-effect SNP in the SNP pair). Error bars denote 95% CIs.

## Notes

### Competing Interest Statement

The authors have declared no competing interest.

### Funding Statement

This research was funded by National Institutes of Health (NIH) grants U01 HG009379, R01 MH101244, R37 MH107649, U01 HG012009 and R01 HG006399. C.C. was funded by the NIGMS 5T32GM007748-44.

### Author Declarations

This research was conducted using the UK Biobank Resource under application #16549 and all data collection was done by the Biobank prior to this project.

